# The Therapeutic Impact of Genetic Evaluation in an Atrial Fibrillation Precision Medicine Clinic

**DOI:** 10.1101/2025.03.28.25324544

**Authors:** J. Lukas Laws, Mahsima Shabani, Hollie L. Williams, Dakota D. Grauherr, Wendy M. Kilbourne, Diane M. Crawford, Isaac Ogunmola, Lili Sun, Zain Virk, Brianna Cathey, Majd A. El-Harasis, Cassady J. Pelphrey, Joseph A. Quintana, Brittany S. Murphy, Giovanni E. Davogustto, M. Edward Ponder, Omeed M. Irani, J. Michael Daw, Bibin T. Varghese, Pablo Saavedra, Robert L. Abraham, Juan C. Estrada, Katherine T. Murray, Walter K. Clair, Sharon T. Shen, Arvindh N. Kanagasundram, Jay A. Montgomery, Christopher R. Ellis, Frank Fish, Travis D. Richardson, George H. Crossley, Rebecca R. Hung, Jeffrey M. Dendy, Adam Wright, Quinn S. Wells, Fei Ye, Harikrishna Tandri, William G. Stevenson, Megan Lancaster, Prince J. Kannankeril, Lynne W. Stevenson, Dan M. Roden, Zachary T. Yoneda, M. Benjamin Shoemaker

## Abstract

**Background and Aims:** Genetic testing is recommended for select patients with atrial fibrillation (AF). The aims of this study were to define the results of genetic evaluation and its therapeutic impact for patients referred to a dedicated AF precision medicine clinic.

**Methods:** Patients diagnosed with AF before age 60 were candidates for referral. In addition to standard evaluation with history, physical exam, and ECG, genetic evaluation included a 3-generation pedigree, cardiac imaging, ambulatory monitoring, and clinical genetic testing with a cardiomyopathy/arrhythmia panel.

**Results:** 264 participants were referred: the median age was 47 years (Q1, Q3: 38, 55), 77 (29%) were female, and 236 (89%) were White. Median age at AF diagnosis was 39 years (Q1, Q3: 31, 48) and median time from AF diagnosis to evaluation was 3.7 years (Q1, Q3: 0.9, 10). 242 patients (92%) underwent genetic testing, which identified a pathogenic or likely pathogenic variant in 48 (20%). The strongest predictors of positive genetic testing were history of cardiomyopathy, infranodal conduction disease, and elevated T1 or late gadolinium enhancement on cardiac MRI (all p<0.05). The strongest predictors of negative genetic testing were obstructive sleep apnea and a normal 12-lead ECG (both p<0.04). Overall, genetic testing changed clinical management in 52% of patients with positive genetic testing, highlighted by 7 new ICD placements and initiation of disease modifying therapy in 16 patients.

**Conclusions:** Genetic testing was positive in 20% of patients with early-onset AF referred to a dedicated AF precision medicine clinic. Genetic testing results changed clinical management in approximately half of genotype-positive patients.

**STRUCTURED GRAPHICAL ABSTRACT:** *Key Question:* Does genetic evaluation of patients with early-onset atrial fibrillation (AF) change their clinical management?

*Key Finding:* Among 246 participants that completed genetic evaluation in a dedicated AF precision medicine clinic, 20% had positive genetic testing with identification of a pathogenic cardiomyopathy or channelopathy variant. These findings led to changes in clinical management in 52% of patients with positive genetic testing.

*Take-home Message:* Genetic evaluation of patients with early-onset AF consists of detailed phenotyping and genetic testing to identify previously undiagnosed genetic disorders. This facilitates earlier diagnosis and clinical intervention. 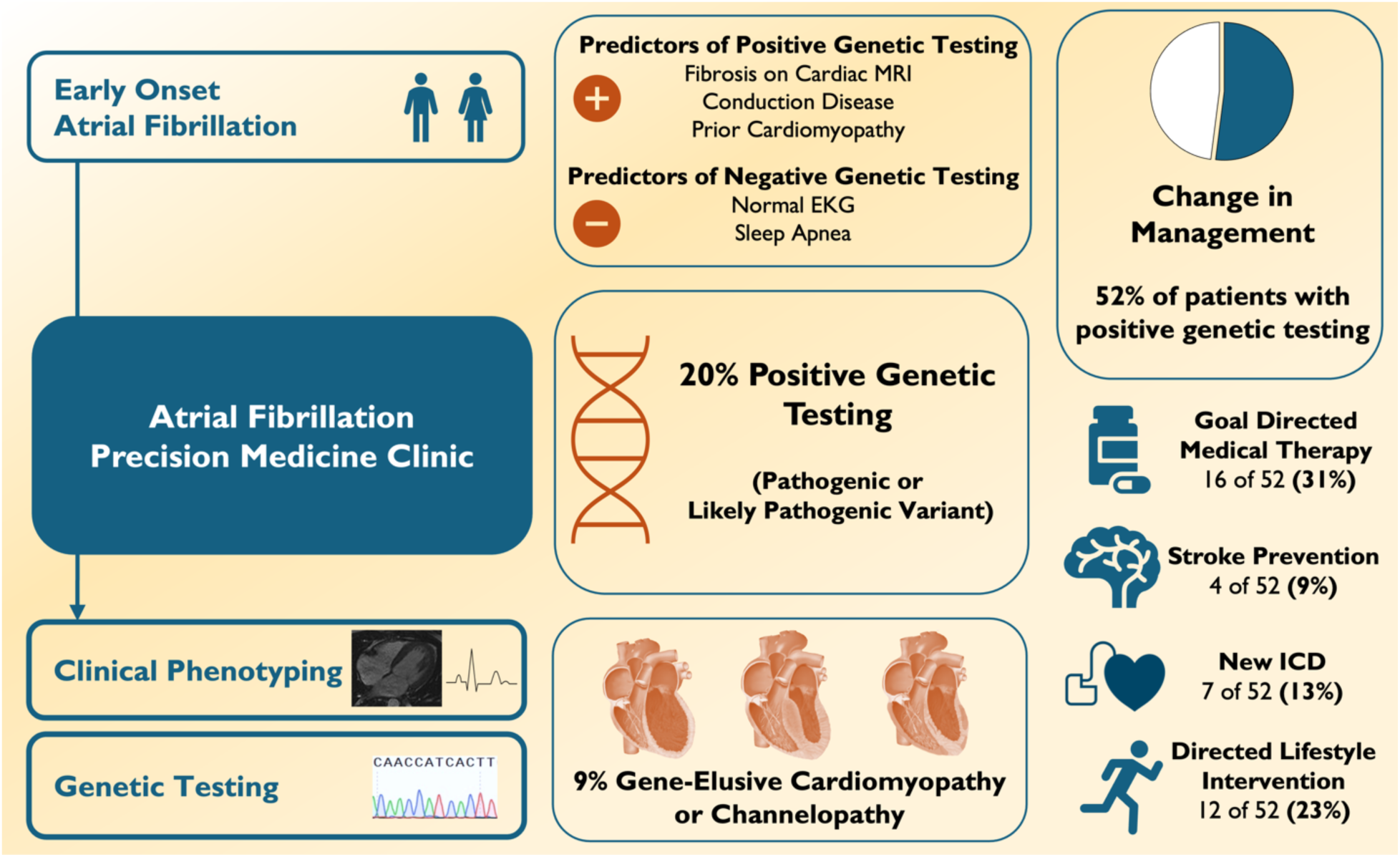

## INTRODUCTION

Atrial fibrillation (AF) is estimated to affect over 30 million people worldwide with most patients diagnosed at an advanced age.(1) AF was not historically considered a genetic syndrome until case reports began emerging of patients and families with early-onset AF and genetic “overlap syndromes”. These patients presented initially with AF and later developed other serious cardiac manifestations such as heart failure, conduction disease, or sudden death due to rare genetic variants. Early examples included syndromes associated with pathogenic variants in *KCNQ1* (Long or Short QT Syndrome), *SCN5A* (Brugada Syndrome, Progressive Cardiac Conduction Disease), and *LMNA* (Arrhythmogenic Cardiomyopathy, ACM).(2–4) At the time, these cases were thought to be extremely rare, so genetic testing was not recommended.(5) That idea was challenged in 2018 with results from large sequencing studies that identified an association between pathogenic variants in *TTN* and early-onset AF.(6,7) *TTN* is the leading monogenic cause of dilated cardiomyopathy (DCM) and pathogenic, truncating variants in *TTN* were found in 6.5% of patients with AF diagnosed before age 30 and 2.1% of patients diagnosed before age 65.(6) These results raised the question of whether clinical genetic testing should be considered for patients with early-onset AF.(8,9) Accordingly, a study in 2021 analyzed 145 genes that were included on clinical cardiomyopathy and arrhythmia gene panels and found that in patients with AF diagnosed before age 65, the prevalence of positive genetic testing (identification of a pathogenic or likely pathogenic variant) was up to 10.1%.(10)

Practice guidelines have incorporated this new evidence and now support the use of genetic testing for select patients with AF.(11,12) The recommendations state that the benefit of genetic testing in AF is to inform prognosis, which is supported by results demonstrating a significantly higher risk of mortality for patients with AF who possess a pathogenic cardiomyopathy or arrhythmia variant.(11–13) The increased mortality risk associated with these variants is likely not from AF directly, but instead from development of an overlapping cardiomyopathy or arrhythmia syndrome that can lead to fatal ventricular arrhythmias and/or heart failure (e.g., ACM, DCM, or Hypertrophic Cardiomyopathy, HCM).(14,15) A major knowledge gap we seek to address is whether genetic testing for patients with early-onset AF can be used to guide therapeutic decision making by facilitating the diagnosis of an AF overlap syndrome.(14) Accordingly, we hypothesized that a detailed evaluation in a dedicated AF precision medicine clinic consisting of genetic testing and detailed phenotyping would have important diagnostic and therapeutic implications for patients with early-onset AF.

## METHODS

### Genetic Evaluation

Patients were referred for genetic evaluation of AF by their clinical provider or by self-referral. Clinical providers could refer based on clinical suspicion for a genetic etiology, or were prompted to refer young patients with AF via an automated Best Practice Alert through the electronic medical record (**Supplemental Figure 1**). “Genetic evaluation” consisted of both genetic testing and clinical phenotyping to evaluate for AF overlap syndromes that may be present with or without positive genetic testing. Patients with cardiomyopathy or ventricular arrhythmias were eligible for enrollment in the registry only if AF was diagnosed prior to cardiomyopathy or if cardiomyopathy was identified concurrently as part of the diagnostic workup of AF. **Figure 1** shows the framework for genetic evaluation. Briefly, patients diagnosed with AF prior to 60 years of age and/or those with a strong family history of AF were candidates for referral based on a suspicion for genetic susceptibility to AF. The initial clinic visit consisted of a comprehensive history and physical, a baseline AF symptom severity questionnaire, 12-lead ECG, construction of a 3-generation pedigree, and genetic counseling. Standard testing included a continuous ambulatory ECG monitor, and cardiac imaging (cardiac MRI or transthoracic echocardiogram). Most patients underwent clinical genetic testing with a comprehensive cardiomyopathy and arrhythmia panel using commercial vendors (Labcorp/Invitae, Burlington, NC; GeneDx, Stamford, CT) or the CLIA-approved Vanderbilt University Medical Center Clinical Genetic Testing Laboratory (Nashville, TN). Some family members underwent targeted genotyping limited to only the familial variant. Depending on the results of the standard clinical evaluation and genetic testing, additional diagnostic testing with an exercise tolerance test, extended ECG monitoring, or a procainamide challenge was performed. Patients were seen for a 3-month follow-up appointment either in-person or by telemedicine to review the results of their tests and develop a longitudinal care plan.

**Figure 1:**
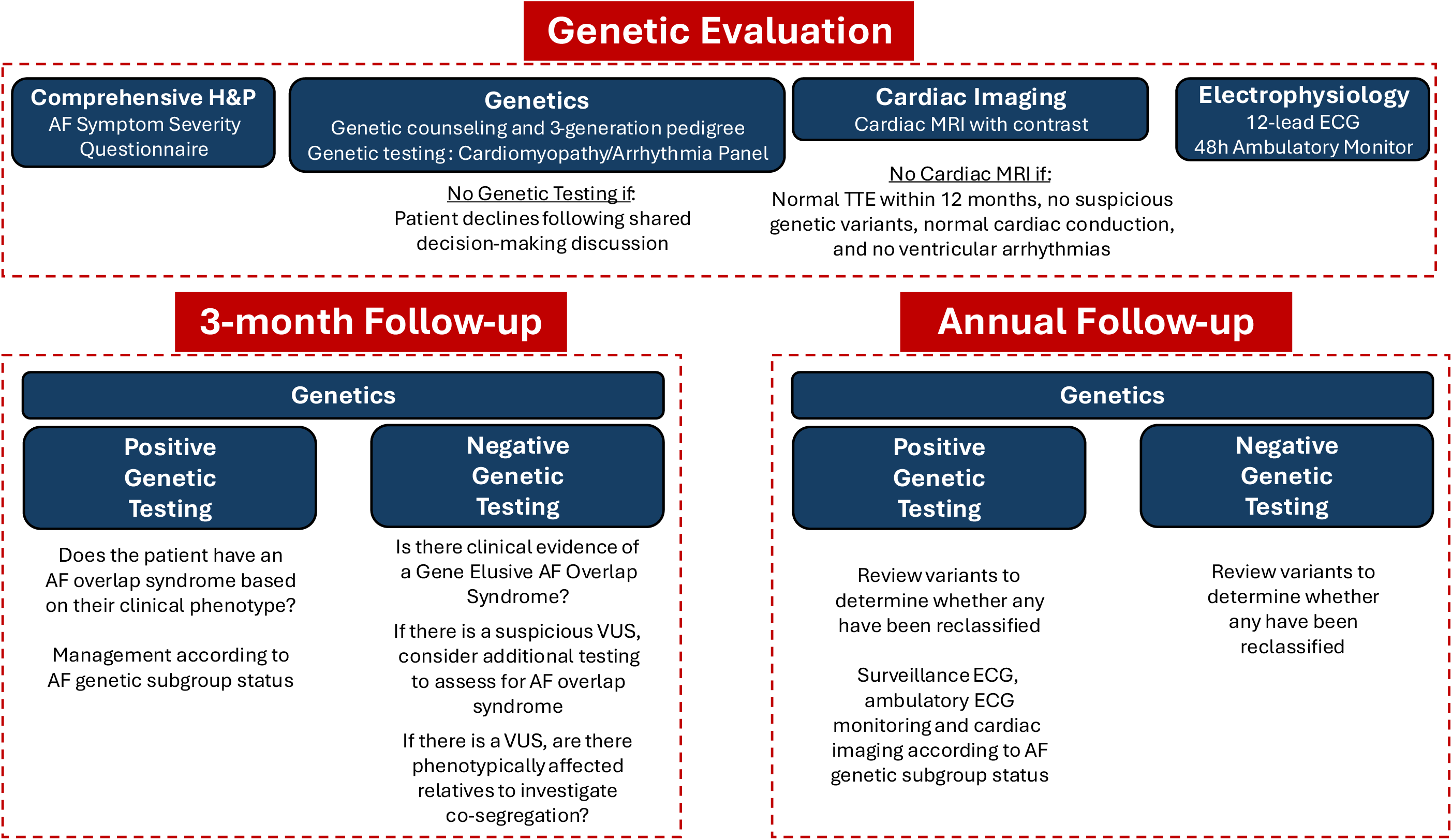
Framework for Evaluation in the Atrial Fibrillation Precision Medicine Clinic. Comprehensive evaluation includes an assessment of both genetic predisposition and clinical phenotyping to evaluate all cardiac manifestations of genetic disease.

### Variant Interpretation

The clinical genetic testing laboratories for this study used the ACMG/AMP (American College of Medical Genetics and Association for Molecular Pathology) Standards and Guidelines for variant interpretation.(16) “Positive” genetic testing was defined as the presence of one or more pathogenic or likely pathogenic (P/LP) variants in a gene with autosomal dominant (AD) inheritance, or two P/LP variants in a gene with autosomal recessive (AR) inheritance. For genes with X-linked inheritance, genetic testing was considered positive for hemizygous men or homozygous women. Due to variable splicing in cardiac and skeletal myocytes, P/LP *TTN* variants were required to be in a cardiac exon with a proportion spliced index (PSI) ≥90% to be classified as positive.(17,18) All patients whose genetic testing results did not meet the “positive” genetic testing criteria were considered to have “negative” genetic testing. Negative genetic testing included those with variants of undetermined significance (VUS), heterozygous carriers of variants in a gene with AR inheritance, and patients with no rare variants reported. Some VUS were designated as “suspicious” if they: 1) possessed characteristics that suggested a potential pathogenic role but lacked sufficient evidence to be definitively classified as pathogenic or likely pathogenic (19), 2) were in a strong/definitive evidence gene, and 3) were potentially consistent with the clinical phenotype of the participant.

### AF Genetic Subgroups

Genes on the cardiomyopathy and arrhythmia genetic testing panel are included because of previously reported association with an inherited cardiomyopathy or arrhythmia syndrome. Genes were classified according to the predominant gene-phenotype association here termed “AF genetic subgroups”. **Table 1** lists the AF genetic subgroups and corresponding genes, and the full gene panel used for evaluation is listed in **Supplemental Table 1**. For this analysis, the AF genetic subgroups are limited to DCM-genes, ACM-genes, HCM-genes, and Channelopathy-genes. While Arrhythmogenic Cardiomyopathy is a broader, non-specific term, it is used here to classify both Arrhythmogenic Right Ventricular Cardiomyopathy (ARVC) and Nondilated Left Ventricular Cardiomyopathy (NDLVC) due to the significant overlap of shared phenotypic features and genetic predisposition between these two diagnoses.(20) Additional detailed methods regarding enrollment, variant interpretation, and phenotype classification are detailed in **Supplemental Methods S.1**.

**TABLE 1:** Major Genes Listed According to Proposed AF Genetic Subgroups.

| DCM | ACM<br>(ARVC, NDLVC) | HCM | Channelopathies<br>(Brugada, LQTS/SQTS, CPVT, PCCD) |
| --- | --- | --- | --- |
| *TTN* | *DSP* | *MYBPC3* | *KCNQ1* |
|  | *EMD (XL)* | *MYH7* | *KCNH2* |
|  | *FLNC* | *ACTC1* | *SCN5A* |
|  | *LMNA* | *ACTN2* | *RYR2* |
|  | *PKP2* | *ALPK3* | *CACNA1C* |
|  | *TRPM4* | *CSRP3* | *CALM1* |
|  | *BAG3* | *MYL2* | *CALM2* |
|  | *DES* | *MYL3* | *CALM3* |
|  | *DSC2* | *PRKAG2* | *CASQ2* |
|  | *DSG2* | *TNNC1* | *HCN4* |
|  | *JUP* | *TNNI3* | *KCNE1* |
|  | *PLN* | *TNNT2* | *KCNJ2* |
|  | *RBM20* | *TPM1* | *TRDN (AR)* |
|  | *TMEM43* |  |  |

### Penetrance of the Ventricular Phenotype and Management Considerations

Patients with AF and positive genetic testing may or may not develop any clinical manifestations of the AF overlap syndrome for which they are genetically susceptible. Official diagnostic criteria defined by practice guidelines were used for DCM, ACM, HCM, Brugada Syndrome (BrS), Long and Short QT Syndrome (LQTS/SQTS), Catecholaminergic Polymorphic VT (CPVT), and Progressive Cardiac Conduction Disease (PCCD) (**Supplemental Table 2**). Patients with AF who also met the diagnostic criteria for an overlapping cardiomyopathy or arrhythmia syndrome were designated as having an “AF-overlap syndrome” with management considerations shown in **Figure 2**. Patients with AF who did not meet diagnostic criteria for their corresponding overlap syndrome were designated as “AF-only”. In this study, all the patients with positive genetic testing could be considered to have penetrant disease with respect to AF, but disease “penetrance” is primarily used to refer to the presence or absence of the corresponding cardiomyopathy or arrhythmia overlap syndrome.

**FIGURE 2:**
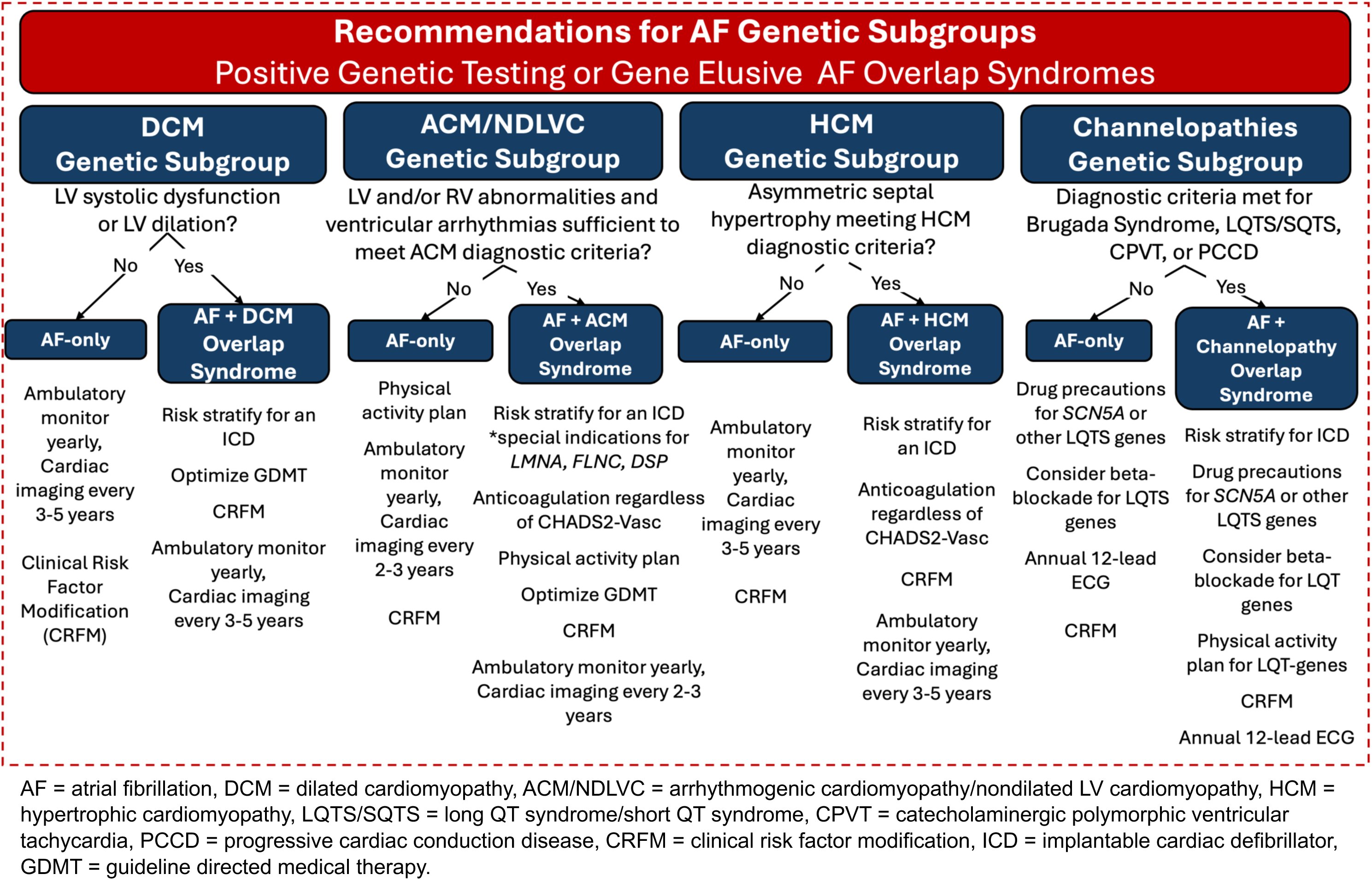
Management Recommendations for the AF Genetic Subgroups. Classification of patients with positive genetic testing using genotype association with risk for AF Overlap Syndromes facilitates individualized surveillance and management changes.

All the syndromes evaluated are also known to occur in the absence of an identifiable genetic variant. For this analysis, the term used is “gene elusive” but elsewhere can also be termed “genotype-negative”. Historically, the rate of gene elusive DCM is estimated to be 50-80%, ACM is 40-50%, HCM is 40%, LQTS is 15-30%, and BrS is 80-90%.(12,21–24) For patients with positive genetic testing or gene elusive AF overlap syndromes, changes in clinical management occurred according to guidelines that exist for the specific overlap syndrome and are presented in **Figure 2** and **Supplemental Table 2.**

### Statistical Analysis

Descriptive statistics for enrollment demographics and characteristics, genetic testing results, and phenotypic evaluation are reported for probands that completed the evaluation (both clinical genetic testing and phenotyping). Penetrance of the ventricular phenotype and changes to clinical management are reported for all patients that completed genetic testing and phenotypic evaluation, including relatives identified through cascade screening and targeted single variant genetic testing. Patients were grouped by positive genetic testing, gene elusive AF-overlap syndrome, or negative genetic evaluation. Univariate analysis of group differences was determined by Chi-square test or Fisher’s exact test for categorical variables, and Kruskal-Wallis or pairwise Wilcoxon rank-sum tests for continuous variables. P-values are reported after correction for multiple testing using false discovery rate, with significance defined as FDR < 0.05. Logistic regression was used to identify potential predictors associated with positive genetic testing, within a multivariable model adjusting for age at AF diagnosis, sex, and time between AF diagnosis and genetic testing to account for age-related penetrance of associated overlap syndromes. All statistical analysis was performed using R statistical software version 4.4.2 (R Foundation for Statistical Computing, Vienna, Austria).

## RESULTS

### Yield of Genetic Testing and Phenotypic Evaluation

Two-hundred-sixty-four patients were referred for genetic evaluation and prospectively enrolled (**Figure 3**). The median age at time of evaluation was 47 years and the median age at AF diagnosis was 39 years. Genetic evaluation was completed in 242 probands (**Table 2)**. Full details of clinical genetic testing are included in **Supplemental Table 3**. Genetic testing was positive in 20% of probands and identified 49 variants (**Figure 4)**, including 1 that was a compound heterozygote (**Table 3)**. Among participants with negative genetic testing, 129 participants (53%) had a VUS and 30 participants (12%) had a VUS that was considered suspicious (**Supplemental Table 4**). Among the remaining participants, 18 participants (7%) were heterozygous carriers of a P/LP variant for an autosomal recessive syndrome and 47 (20% had no rare variants reported). Among the participants with negative genetic testing, clinical testing results identified a gene elusive AF overlap syndrome in 22 (9%) (**Figure 3**).

**Figure 3:**
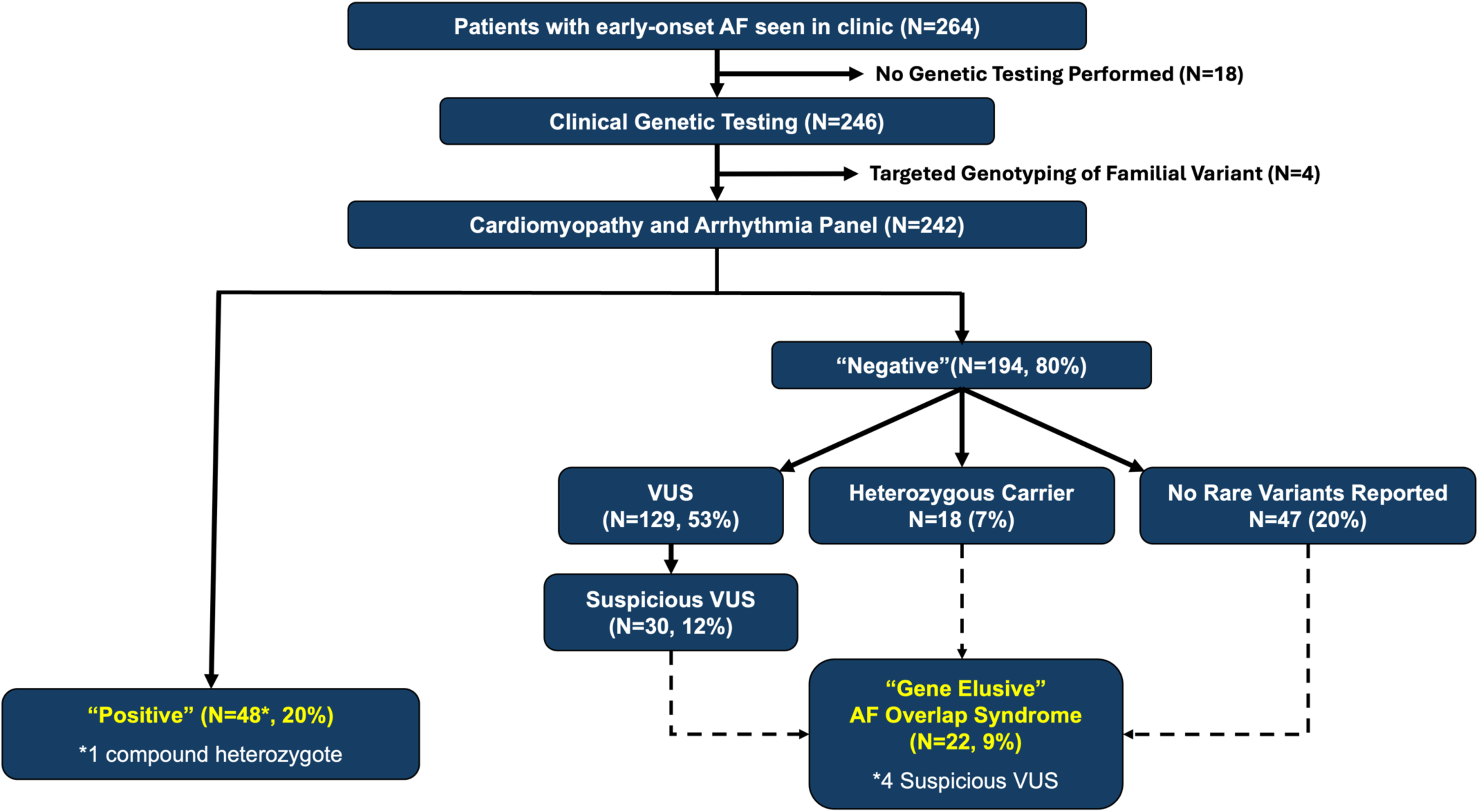
Genetic Testing Results from the Atrial Fibrillation Precision Medicine Clinic.

**FIGURE 4:**
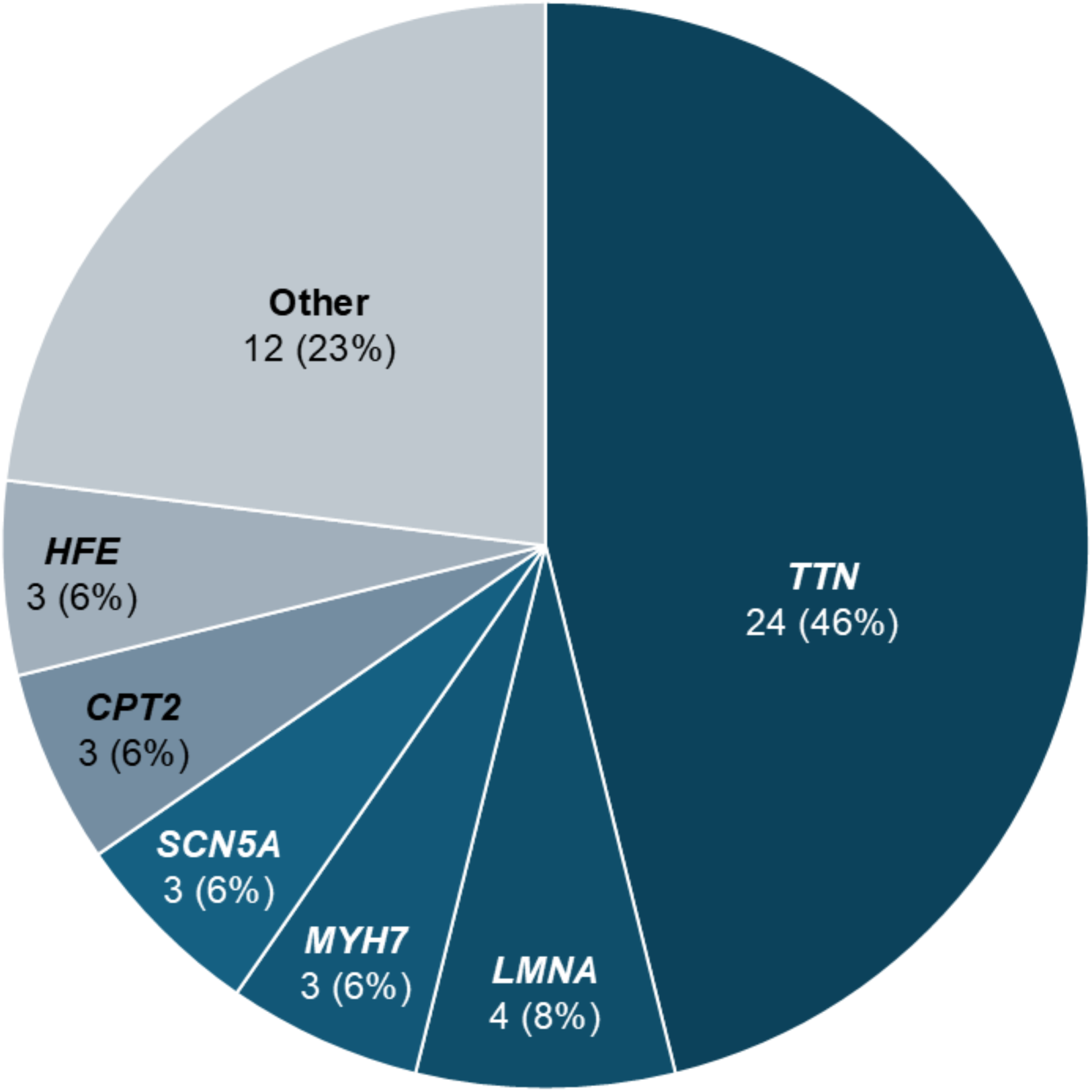
Positive genetic testing results according to gene. Pathogenic or likely pathogenic *TTN* variants were the most prevalent of patients with positive genetic testing. Other genes with strong association with cardiomyopathy *LMNA* and *MYH7* were also prevalent, in addition to the channelopathy gene *SCN5A* associated with Brugada syndrome. Variants in *LMNA* and *MYH7* were the other genes that comprised the largest individual patient pools, with other individual genes less commonly discovered in this cohort.

**FIGURE 5:**
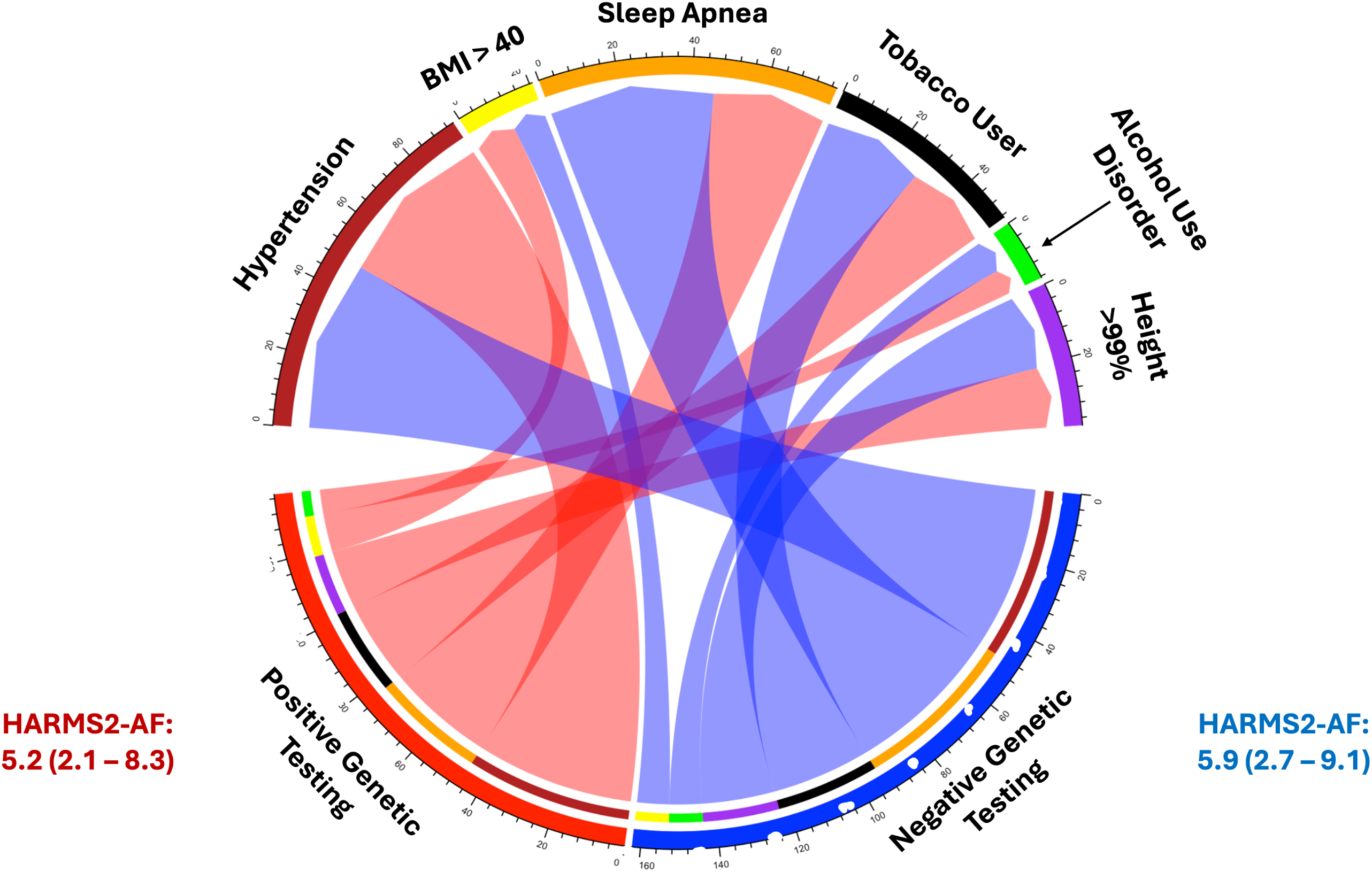
Chord diagram illustrates patients with major clinical risk factors for AF have positive genetic testing. HARMS2-AF is a clinical risk score that found no difference in aggregated clinical risk estimates between patients with positive versus negative genetic testing (p = 0.76).

**Table 2:**
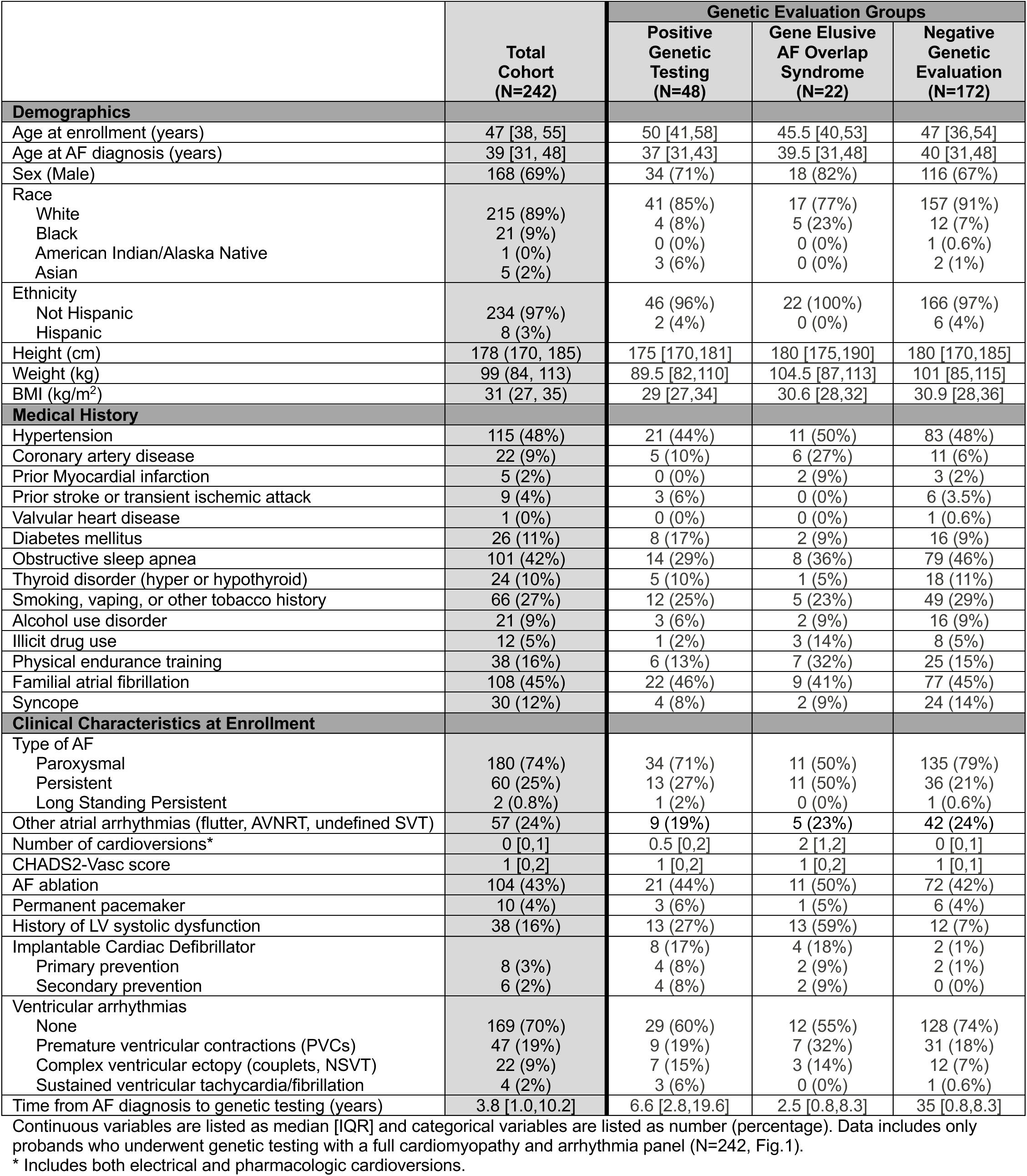
Baseline demographics and clinical characteristics of patients at time of referral to the AF Precision Medicine Clinic.

**TABLE 3:** Variants in probands with positive genetic testing.

| Patient | Gene | Inheritance Pattern | Predominant Overlap Syndrome for Gene | c.DNA | p. Amino Acid | Variant Class |
| --- | --- | --- | --- | --- | --- | --- |
| 1 | *TTN* | AD | DCM | c.56572C>T | p.Arg18858* | Likely pathogenic |
| 2 | *TTN* | AD | DCM | c.3642T>A | p.Tyr1214* | Likely pathogenic |
| 3 | *TTN* | AD | DCM | c.70162C>T | p.Arg23388* | Pathogenic |
| 4 | *TTN* | AD | DCM | c.100587G>A | p.Trp33529* | Likely pathogenic |
| 5 | *TTN* | AD | DCM | c.12587C>A | p.Ser4196* | Likely pathogenic |
| 6 | *TTN* | AD | DCM | c.56572C>T | p.Arg18858* | Likely pathogenic |
| 7 | *TTN* | AD | DCM | c.45316_45320dup | p.Arg15108Alafs*71 | Pathogenic |
| 8 | *TTN* | AD | DCM | c.98299_98300del | p.Arg32767Glyfs*2 | Likely pathogenic |
| 9 | *TTN* | AD | DCM | c.73031G>A | p.Trp24344* | Likely pathogenic |
| 10 | *TTN* | AD | DCM | c.9448C>T | p.Arg3150* | Likely pathogenic |
| 11 | *TTN* | AD | DCM | c.54120del | p.Gly18041Alafs*44 | Likely pathogenic |
| 12 | *TTN* | AD | DCM | c.94263del | p.Ala31422Profs*35 | Likely pathogenic |
| 13 | *TTN* | AD | DCM | c.67095_67908dupTCTT | p.Thr22637Serfs*3 | Likely pathogenic |
| 14 | *TTN* | AD | DCM | c.67833C>G | p.Tyr22611* | Likely pathogenic |
| 15 | *TTN* | AD | DCM | c.53599G>T | p.Glu17867* | Likely pathogenic |
| 16 | *TTN* | AD | DCM | c.107223+2T>C | Intronic | Likely pathogenic |
| 17 | *TTN* | AD | DCM | c.42014_42021del | p.Arg14005Thrfs*4 | Likely pathogenic |
| 18 | *TTN* | AD | DCM | c.49870C>T | p.Arg16624* | Likely pathogenic |
| 19 | *TTN* | AD | DCM | c.58732+2T>C | Intronic | Likely pathogenic |
| 20 | *TTN* | AD | DCM | c.77145dup | p.Ser25716Leufs*8 | Likely pathogenic |
| 21 | *TTN* | AD | DCM | c.57603C>A | p.Cys19201* | Likely pathogenic |
| 22 | *TTN* | AD | DCM | c.48167del | p.Pro16056Glnfs*7 | Likely pathogenic |
| 23 | *TTN* | AD | DCM | c.63370C>T | p.Gln21124* | Likely pathogenic |
| 24 | *LMNA* | AD | ACM | c.658C>T | p.Arg220Cys | Likely pathogenic |
| 25 | *LMNA* | AD | ACM | c.647G>A | p.Arg216His | Likely pathogenic |
| 26 | *LMNA* | AD | ACM | c.1146C>T | p.Gly382= | Likely pathogenic |
| 27 | *PKP2* | AD | ACM | c.1237C>T | p.Arg413* | Pathogenic |
| 28 | *FLNC* | AD | ACM | c.970-4A>G | Intronic | Pathogenic |
| 29 | *EMD* | AD | ACM | Complete Deletion | Complete Deletion | Pathogenic |
| 30 | *TRPM4* | AD | ACM | c.3119T>C | p.Iso1040Thr | Pathogenic |
| 31* | *DSP* | AD | ACM | c.5212C>T | p.Arg1738* | Pathogenic |
| 32 | *MYH7* | AD | HCM | c.4348G>A | p.Asp1450Asn | Pathogenic |
| 33 | *MYH7* | AD | HCM | c.1988G>A | p.Arg663His | Pathogenic |
| 34 | *MYBPC3* | AD | HCM | c.833delG | p.Gly278Glufs*22 | Pathogenic |
| 35 | *SCN5A* | AD | Channelopathy | c.2369T>A | p.Ile790Asn | Likely pathogenic |
| 36 | *SCN5A* | AD | Channelopathy | c.393-2A>G | Intronic | Likely pathogenic |
| 37 | *SCN5A* | AD | Channelopathy | c.5126C>T | p.Thr1709Met | Likely pathogenic |
| 38 | *KCNQ1* | AD | Channelopathy | c.564G>A | p.Trp188* | Pathogenic |
| 31* | *HFE* | AR | Hemochromatosis | c.845G>A/ c.187C>G | p.Cys282Tyr/p.His63Asp | Pathogenic |
| 39 | *HFE* | AR | Hemochromatosis | c.845G>A/ c.187C>G | p.Cys282Tyr/p.His63Asp | Pathogenic |
| 40 | *HFE* | AR | Hemochromatosis | c.845G>A/ c.187C>G | p.Cys282Tyr/p.His63Asp | Pathogenic |
| 41 | *HFE* | AR | Hemochromatosis | c.845G>A/ c.187C>G | p.Cys282Tyr/p.His63Asp | Pathogenic |
| 42 | *CPT2* | AD/AR | CPT2 Deficiency | c.370C>T | p.Arg124* | Pathogenic |
| 43 | *CPT2* | AD/AR | CPT2 Deficiency | c.691C>T | p.Arg231Trp | Likely pathogenic |
| 44 | *CPT2* | AD/AR | CPT2 Deficiency | c.338C>T | p.Ser113Leu | Pathogenic |
| 45 | *TTR* | AD | TTR Amyloidosis | c.424G>A | p.Val142Ile | Pathogenic |
| 46 | *TTR* | AD | TTR Amyloidosis | c.424G>A | p.Val142Ile | Pathogenic |
| 47 | *LZTR1* | AD/AR | Noonan Syndrome | c.485G>A | p.Trp162* | Pathogenic |
| 48 | *GJA5* | AD | Familial AF | Complete Deletion | Complete Deletion | Pathogenic |

**TABLE 4:** Participants with Gene Elusive AF Overlap Syndromes (N=22). All were diagnosed with AF prior to the development of a cardiomyopathy or channelopathy AF-overlap syndrome.

|  | Diagnosis | Age Range at AF Diagnosis | Ventricular Structure and Function | Ventricular Arrhythmias | Sinus Node and Conduction | Family History | Genetic Testing |
| --- | --- | --- | --- | --- | --- | --- | --- |
| 1 | ACM/NDLVC | 56-60 | Non-dilated LV with hypokinesis<br>Worst LVEF 10%, now 50%<br>Septal intramural LGE | Non-sustained VT.<br>Negative EP Study | SSS requiring PPM,<br>LBBB | Negative | Negative,<br>Suspicious VUS in<br>*ACTC1* c.309C>A |
| 2 | ACM/NDLVC | 21-25 | Non-dilated LV with normal function<br>RV dilation & hypokinesis (RVEF 40%)<br>No LGE | Non-sustained VT | Normal | SCD<br>ICD | Negative,<br>VUS x 1 |
| 3 | ACM/NDLVC | 36-40 | Non-dilated chambers with biventricular<br>systolic dysfunction (LVEF 38%) | Non-sustained VT | Normal | SCD | Negative,<br>No rare variants |
| 4 | ACM/NDLVC | 31-35 | Non-dilated LV/RV with hypokinesis<br>LVEF 47%<br>Ring-like intramural LGE | PVCs > 500/day | IVCD | Familial AF | Negative,<br>VUS x 1 |
| 5 | ACM/NDLVC | 56-60 | Non-dilated LV with hypokinesis<br>Worst LVEF 30%, now 65%<br>MRI uninterpretable | Ventricular couplets | Normal | Familial AF,<br>Cardiomyopathy | Negative,<br>VUS x 1 |
| 6 | ACM/NDLVC | 46-50 | Non-dilated LV with hypokinesis<br>Worst LVEF 40%, now 57%<br>No LGE | VT/VF arrest,<br>Secondary<br>prevention ICD | IVCD | Negative | Negative,<br>Suspicious VUS in<br>*TRPM4* c.2295dup |
| 7 | ACM/NDLVC | 41-45 | Non-dilated LV with hypokinesis<br>Worst LVEF 15%, now 40%<br>Ring-like intramural LGE | Non-sustained VT | Bifascicular block | Cardiomyopathy | Negative,<br>VUS x 2 |
| 8 | ACM/NDLVC | 51-55 | Non-dilated LV with hypokinesis<br>LVEF now 46%<br>Unable to assess LGE | PVCs > 2500/day,<br>Non-sustained VT,<br>Primary Prevention<br>ICD for syncope | Normal | Familial AF,<br>Cardiomyopathy,<br>SCD | Negative,<br>No rare variants |
| 9 | ACM/NDLVC | 46-50 | Non-dilated LV with hypokinesis<br>LVEF 35%<br>No LGE. Elevated T2 | PVCs > 9000/day<br>Non-sustained VT | Normal | Familial AF | Negative,<br>VUS x 2 |
| 10 | DCM | 31-35 | Dilated LV with hypokinesis<br>Worst LVEF 42%, now 48%<br>No LGE | Negligible | Normal | SCD | Negative,<br>VUS x 1 |
| 11 | DCM | 16-20 | Dilated LV with hypokinesis<br>Worst LVEF 42%, now 50%<br>No LGE | PVCs >1000/day,<br>Ventricular couplets | Normal | Negative | Negative,<br>Suspicious VUS in<br>*TTN* c.58870G>A |
| 12 | DCM | 31-35 | Dilated LV with hypokinesis<br>Worst LVEF 21%, now 60%<br>No LGE, but elevated T1/T2 | Negligible | Normal | Familial AF | Negative,<br>VUS x 1 |
| 13 | DCM | 36-40 | Non-dilated LV with hypokinesis<br>Worst LVEF 28%, now 55%<br>No LGE | Negligible | Normal | Familial AF | Negative,<br>VUS x 4 |
| 14 | DCM | 56-60 | Dilated LV with hypokinesis<br>Worst LVEF 20%, now 50%<br>Septal intramural LGE | Negligible | Normal | Familial AF,<br>Cardiomyopathy with<br>heart transplant | Negative,<br>No rare variants |
| 15 | DCM | 36-40 | Dilated LV/RV with hypokinesis<br>Worst LVEF 10%, now 65%<br>No LGE | Negligible | Normal | Negative | Negative,<br>Suspicious VUS in<br>SCN5A c.5701 G>A |
| 16 | DCM | 46-50 | Dilated LV with hypokinesis<br>Worst LVEF 10%, now 60%<br>Unable to obtain cMRI | Negligible | Normal | Negative | Negative,<br>VUS x 2 |
| 17 | DCM | 26-30 | Non-dilated LV with hypokinesis<br>Worst LVEF 35%, now 45%<br>Unable to obtain cMRI | PVCs > 3000/day,<br>Ventricular couplets | Normal | Negative | Negative,<br>Heterozygous<br>carrier status |
| 18 | DCM | 36-40 | Dilated LV with hypokinesis<br>Worst LVEF 15%, now 50%<br>No LGE | Negligible | Normal | Familial AF | Negative,<br>Heterozygous<br>carrier status |
| 19 | HCM | 26-30 | Asymmetric LVH<br>LV septum 1.5 cm, posterior wall 1.2 cm<br>No LGE, elevated T2 | Negligible | Normal | Negative | Negative,<br>Heterozygous<br>carrier status |
| 20 | HCM | 41-45 | Asymmetric LVH<br>LV septum 1.5 cm, posterior wall 1.1 cm<br>Septal intramural LGE, elevated T2 | Negligible | Normal | HCM<br>SCD | Negative,<br>No rare variants |
| 21 | Idiopathic VF | 26-30 | Normal biventricular size and function<br>Myxomatous mitral valve with bileaflet<br>prolapse and trace MR | VT/VF arrest,<br>Secondary<br>prevention ICD | Normal | Negative | Negative,<br>VUS x 1 |
| 22 | PCCD | 41-45 | Normal biventricular size and function | Negligible | CHB requiring PPM | Negative | Negative,<br>VUS x 1 |

All participants underwent a 12-lead ECG, 209 (85%) ambulatory monitoring, and 230 (93%) cardiac imaging according to the framework in **Figure 1**. Additional testing was performed in 112 participants (46%) with exercise treadmill ECG, and 6 (2%) with a sodium channel blocker (procainamide) challenge. Findings from phenotypic evaluation including ECG, ambulatory ECG monitoring, and cardiac imaging are shown in **Table 5** and **Supplemental Table 5**.

**TABLE 5:**
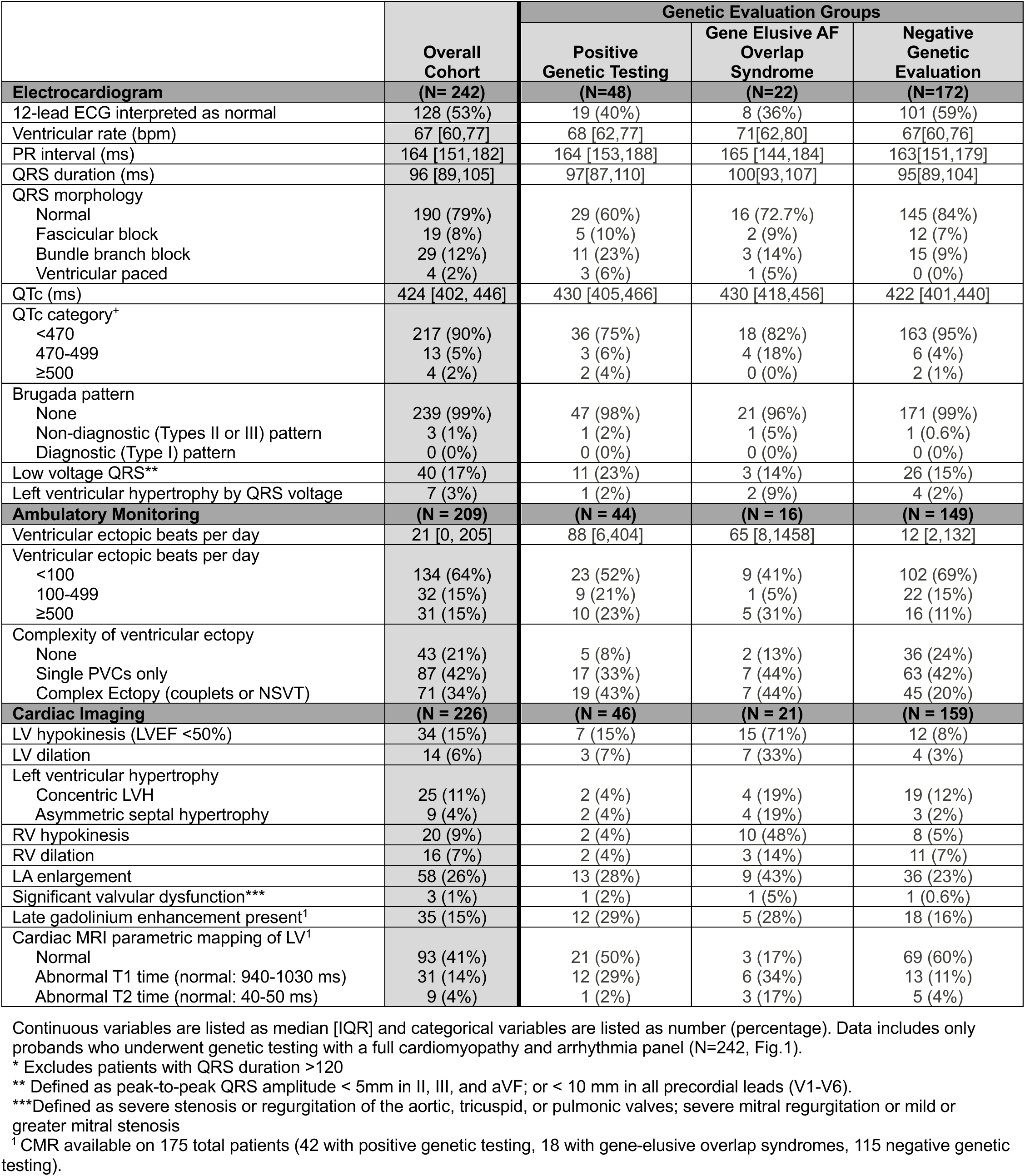
Electrocardiogram, Cardiac Imaging, and Ambulatory Monitoring Data.

|  | Overall Cohort | Genetic Evaluation Groups |  |  |
| --- | --- | --- | --- | --- |
|  |  | Positive Genetic Testing | Gene Elusive AF Overlap Syndrome | Negative Genetic Evaluation |
| **Electrocardiogram** | **(N= 242)** | **(N=48)** | **(N=22)** | **(N=172)** |
| 12-lead ECG interpreted as normal | 128 (53%) | 19 (40%) | 8 (36%) | 101 (59%) |
| Ventricular rate (bpm) | 67 [60,77] | 68 [62,77] | 71[62,80] | 67[60,76] |
| PR interval (ms) | 164 [151,182] | 164 [153,188] | 165 [144,184] | 163[151,179] |
| QRS duration (ms) | 96 [89,105] | 97[87,110] | 100[93,107] | 95[89,104] |
| QRS morphology |  |  |  |  |
| Normal | 190 (79%) | 29 (60%) | 16 (72.7%) | 145 (84%) |
| Fascicular block | 19 (8%) | 5 (10%) | 2 (9%) | 12 (7%) |
| Bundle branch block | 29 (12%) | 11 (23%) | 3 (14%) | 15 (9%) |
| Ventricular paced | 4 (2%) | 3 (6%) | 1 (5%) | 0 (0%) |
| QTc (ms) | 424 [402, 446] | 430 [405,466] | 430 [418,456] | 422 [401,440] |
| QTc category<sup>+</sup> |  |  |  |  |
| <470 | 217 (90%) | 36 (75%) | 18 (82%) | 163 (95%) |
| 470-499 | 13 (5%) | 3 (6%) | 4 (18%) | 6 (4%) |
| ≥500 | 4 (2%) | 2 (4%) | 0 (0%) | 2 (1%) |
| Brugada pattern |  |  |  |  |
| None | 239 (99%) | 47 (98%) | 21 (96%) | 171 (99%) |
| Non-diagnostic (Types II or III) pattern | 3 (1%) | 1 (2%) | 1 (5%) | 1 (0.6%) |
| Diagnostic (Type I) pattern | 0 (0%) | 0 (0%) | 0 (0%) | 0 (0%) |
| Low voltage QRS<sup>**</sup> | 40 (17%) | 11 (23%) | 3 (14%) | 26 (15%) |
| Left ventricular hypertrophy by QRS voltage | 7 (3%) | 1 (2%) | 2 (9%) | 4 (2%) |
| **Ambulatory Monitoring** | **(N = 209)** | **(N = 44)** | **(N = 16)** | **(N = 149)** |
| Ventricular ectopic beats per day | 21 [0, 205] | 88 [6,404] | 65 [8,1458] | 12 [2,132] |
| Ventricular ectopic beats per day |  |  |  |  |
| <100 | 134 (64%) | 23 (52%) | 9 (41%) | 102 (69%) |
| 100-499 | 32 (15%) | 9 (21%) | 1 (5%) | 22 (15%) |
| ≥500 | 31 (15%) | 10 (23%) | 5 (31%) | 16 (11%) |
| Complexity of ventricular ectopy |  |  |  |  |
| None | 43 (21%) | 5 (8%) | 2 (13%) | 36 (24%) |
| Single PVCs only | 87 (42%) | 17 (33%) | 7 (44%) | 63 (42%) |
| Complex Ectopy (couplets or NSVT) | 71 (34%) | 19 (43%) | 7 (44%) | 45 (20%) |
| **Cardiac Imaging** | **(N = 226)** | **(N = 46)** | **(N = 21)** | **(N = 159)** |
| LV hypokinesis (LVEF <50%) | 34 (15%) | 7 (15%) | 15 (71%) | 12 (8%) |
| LV dilation | 14 (6%) | 3 (7%) | 7 (33%) | 4 (3%) |
| Left ventricular hypertrophy |  |  |  |  |
| Concentric LVH | 25 (11%) | 2 (4%) | 4 (19%) | 19 (12%) |
| Asymmetric septal hypertrophy | 9 (4%) | 2 (4%) | 4 (19%) | 3 (2%) |
| RV hypokinesis | 20 (9%) | 2 (4%) | 10 (48%) | 8 (5%) |
| RV dilation | 16 (7%) | 2 (4%) | 3 (14%) | 11 (7%) |
| LA enlargement | 58 (26%) | 13 (28%) | 9 (43%) | 36 (23%) |
| Significant valvular dysfunction<sup>***</sup> | 3 (1%) | 1 (2%) | 1 (5%) | 1 (0.6%) |
| Late gadolinium enhancement present<sup>1</sup> | 35 (15%) | 12 (29%) | 5 (28%) | 18 (16%) |
| Cardiac MRI parametric mapping of LV<sup>1</sup> |  |  |  |  |
| Normal | 93 (41%) | 21 (50%) | 3 (17%) | 69 (60%) |
| Abnormal T1 time (normal: 940-1030 ms) | 31 (14%) | 12 (29%) | 6 (34%) | 13 (11%) |
| Abnormal T2 time (normal: 40-50 ms) | 9 (4%) | 1 (2%) | 3 (17%) | 5 (4%) |

<sup>+</sup> Excludes patients with QRS duration >120
<sup>\*\*</sup> Defined as peak-to-peak QRS amplitude < 5mm in II, III, and aVF; or < 10 mm in all precordial leads (V1-V6).
<sup>\*\*\*</sup>Defined as severe stenosis or regurgitation of the aortic, tricuspid, or pulmonic valves; severe mitral regurgitation or mild or greater mitral stenosis
<sup>1</sup> CMR available on 175 total patients (42 with positive genetic testing, 18 with gene-elusive overlap syndromes, 115 negative genetic testing).

### Predictors of Positive Genetic Testing During Initial Evaluation

Multivariable logistic regression models were used to estimate the association between specific demographics and clinical characteristics and the likelihood of having positive genetic testing, adjusting for age at AF diagnosis, sex, and time between AF onset and clinical genetic testing (**Figure 6A**). From the clinical history, a previous diagnosis of cardiomyopathy (OR 5.3, 95% CI 2.1-13.3, p <0.001) was significantly associated with positive genetic testing. Family history of AF, type of AF (paroxysmal, persistent, permanent), and number of cardioversions were not significantly associated with positive genetic testing. Obstructive sleep apnea was the only clinical risk factor for AF that predicted negative genetic testing (OR 0.4, 95% CI 0.2-0.8, p = 0.013). The likelihood of a positive genetic test tends to be lower for patients with other clinical risk factors such as hypertension, height >99^th^ percentile, and alcohol use disorder that did not meet threshold for statistical significance. Tobacco use (OR 0.9, 95% CI 0.4-1.8) and obesity (BMI 30-39, OR 1.0, 95% CI 0.3-3.0; BMI>40, OR 1.9, 95% CI 0.5-7.3) had minimal estimated effect for positive genetic testing in the model.

**Figure 6:**
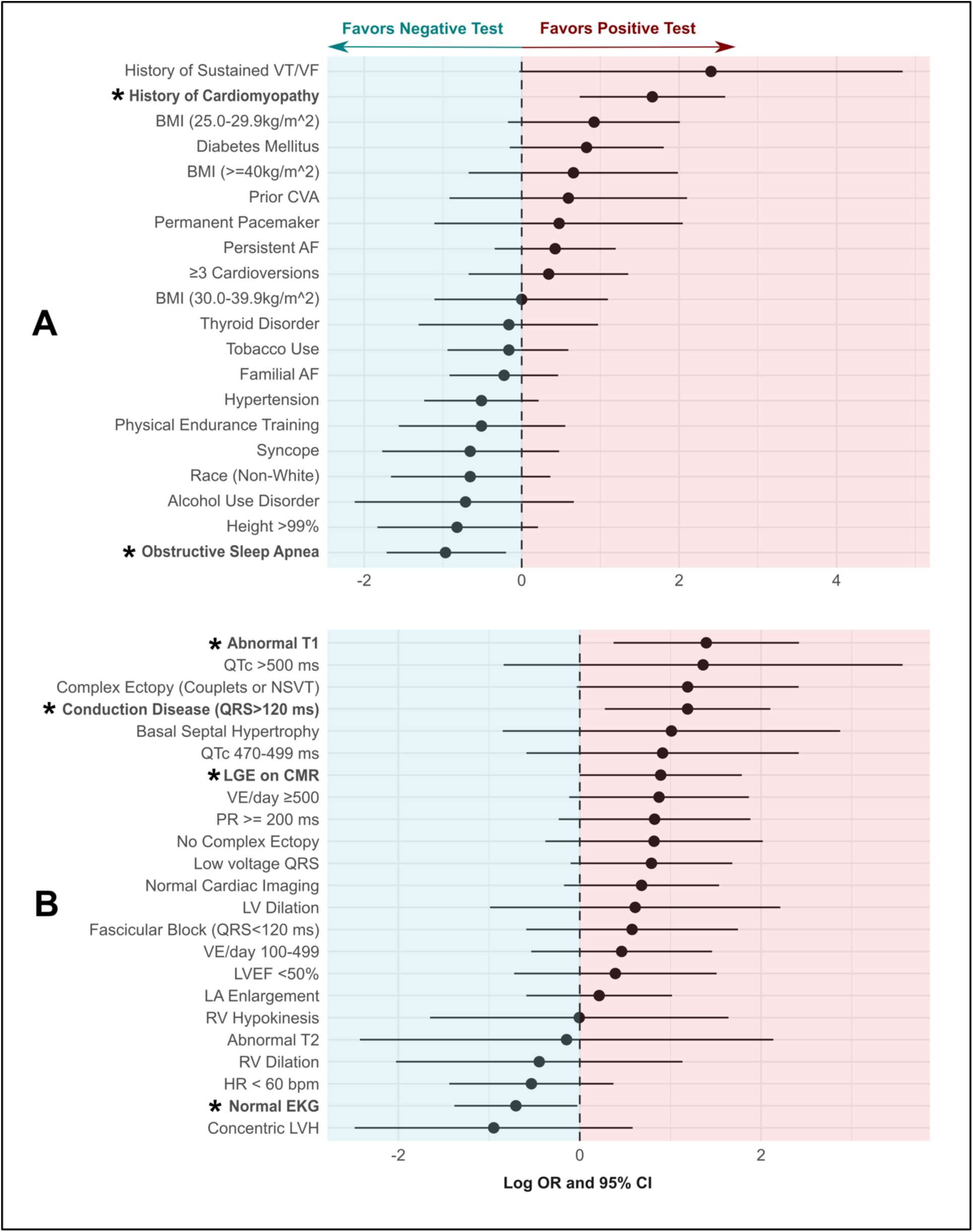
Forest plot of clinical and historical data (Panel A) and ECG, cardiac imaging, and ambulatory monitoring data (Panel B) and the association with positive genetic testing. The analysis is adjusted for age at AF diagnosis, sex, and time between AF diagnosis and clinical genetic testing. Patients with gene-elusive AF overlap syndromes were excluded from the prediction models.

### Predictors of Positive Genetic Testing from Electrocardiographic and Imaging Evaluation

Results from clinical phenotyping were also assessed in a multivariable model adjusted for age at AF diagnosis, sex, and time from AF diagnosis to clinical genetic testing (**Figure 6B**). Findings from cardiac MRI were strongly associated with positive genetic testing, as the strongest predictor was elevated T1 time (OR 4.0, 95% CI 1.5-11.2, p = 0.007). Late gadolinium enhancement (LGE) from CMR was also associated with a positive genetic test (OR 2.4, 95% CI 1.0-6.0, p = 0.050). From the initial ECG, infranodal conduction disease (RBBB/LBBB/IVCD) was the only statistically significant predictor of positive genetic testing (OR 3.3, 95% CI 1.3-8.2, p = 0.011), while a 12-lead ECG at time of presentation interpreted as “normal” was associated with significantly lower likelihood of a positive genetic evaluation (OR 0.5, 95% CI 0.3-1.0, p = 0.042). Complex and frequent ventricular ectopy recorded on ambulatory monitoring was associated with increased likelihood of positive genetic testing, though this result did not reach statistical significance (OR 3.3, 95% CI 0.9-11.2, p = 0.057).

### Penetrance of the Ventricular Phenotype in Participants with Positive Genetic Testing

After completion of genetic testing and phenotypic evaluation in probands and gene-positive family members (N = 246), penetrance of the ventricular phenotype in genotype-positive participants was assessed according to the predominant phenotype association shown in **Table 1**. Participants harboring a variant in genes associated with ACM had the highest penetrance of the ventricular phenotype with 89% (8/9) found to have ACM as shown in **Figure 7**. While DCM-associated variants (including all P/LP *TTN* variants) were the most common result of positive genetic testing (all *TTN* in this cohort), 54% (13/24) of these participants had a DCM phenotype. Genes associated with HCM (N = 5) and Channelopathies (N = 4) were identified less often but had similar penetrance of the ventricular phenotype with 40% of P/LP HCM variant carriers with overt phenotypic expression, and 50% of P/LP channelopathy participants found to have LQTS or Brugada Syndrome.

**FIGURE 7:**
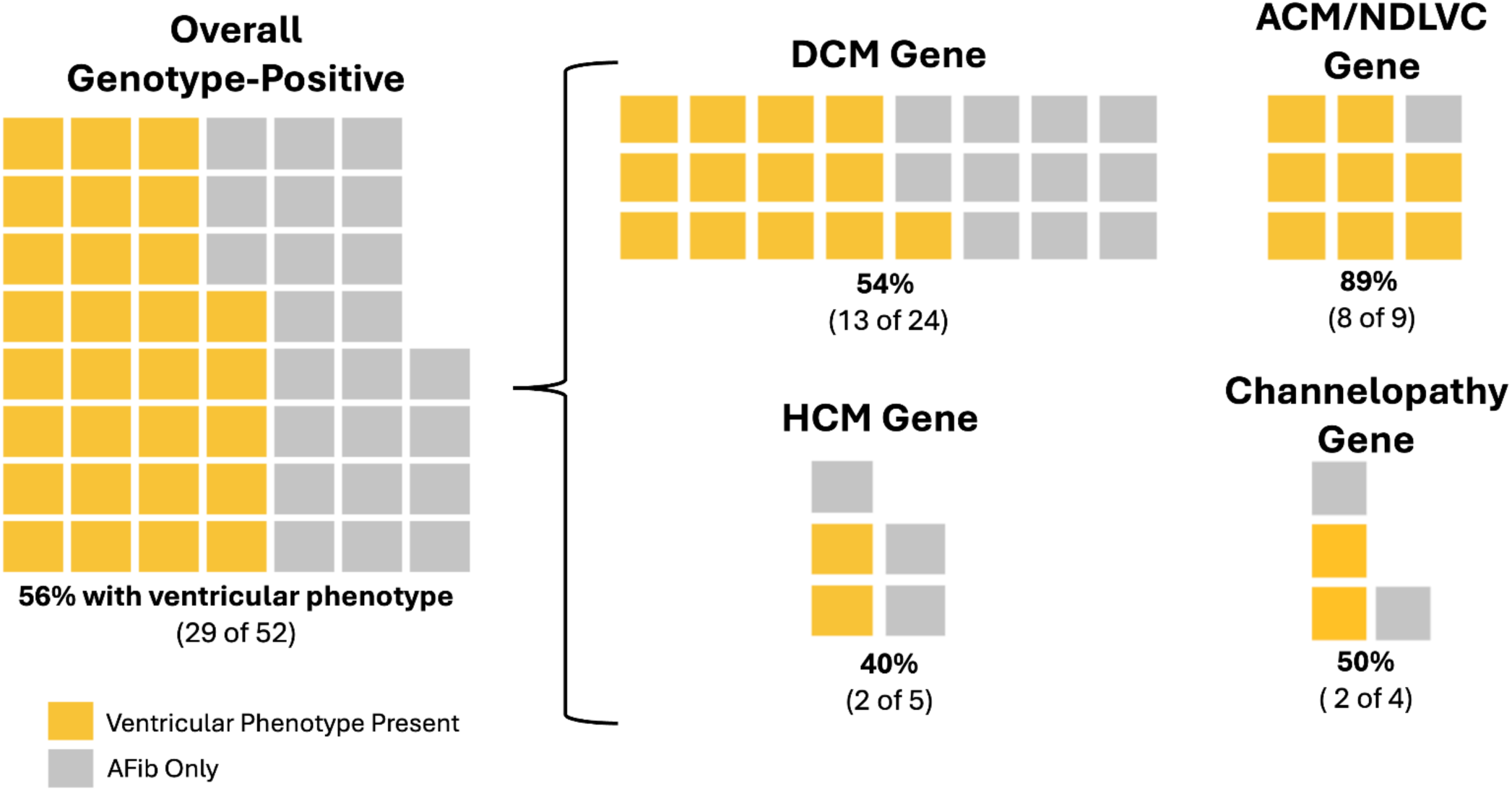
Penetrance of the ventricular phenotype. Each box represents one patient with positive genetic testing. The penetrance of the ventricular phenotype was 56% in the overall cohort. When stratified according to specific AF genetic subgroups, penetrance of the ventricular phenotype ranged from 40% to 89%.

### Changes to Clinical Management

Impact on clinical care was assessed prospectively based on interventions performed after completion of the genetic and phenotypic evaluation. Complete details on changes to clinical management are presented in **Supplemental Table 6**. Participants with positive genetic testing had a change to management in 52% (27/52) of cases, and management was impacted in 11% (7/22) of participants with Gene Elusive AF Overlap Syndromes **(Figure 8A)**. The frequency and type of clinical management was also different among the AF Genetic Subgroups with a phenotype present (**Figure 8B).** 92% of participants with an ACM phenotype had a change to clinical management, most commonly initiation of guideline-directed medical therapy (GDMT) followed by recommendations for directed lifestyle intervention, such as exercise precautions for participants with desmosomal variants. Changes to clinical management were made less often (9 of 27, 33%) in participants with an identified DCM phenotype. Notably, all participants with HCM variants or phenotype had a change to clinical management. Results of the genetic evaluation facilitated early intervention to mitigate the risk of sudden cardiac death, with implantation of a new ICD in 7 participants. For participants with channelopathy variants or phenotype, directed lifestyle interventions regarding medication use and precautions were the most common management change. Specifically, 1 participant had a pathogenic *KCNQ1* variant and was given precautions against QT-prolonging medications and 3 participants had a pathogenic *SCN5A* variant and were given precautions against sodium channel blocking medications and fever precautions.

**Figure 8:**
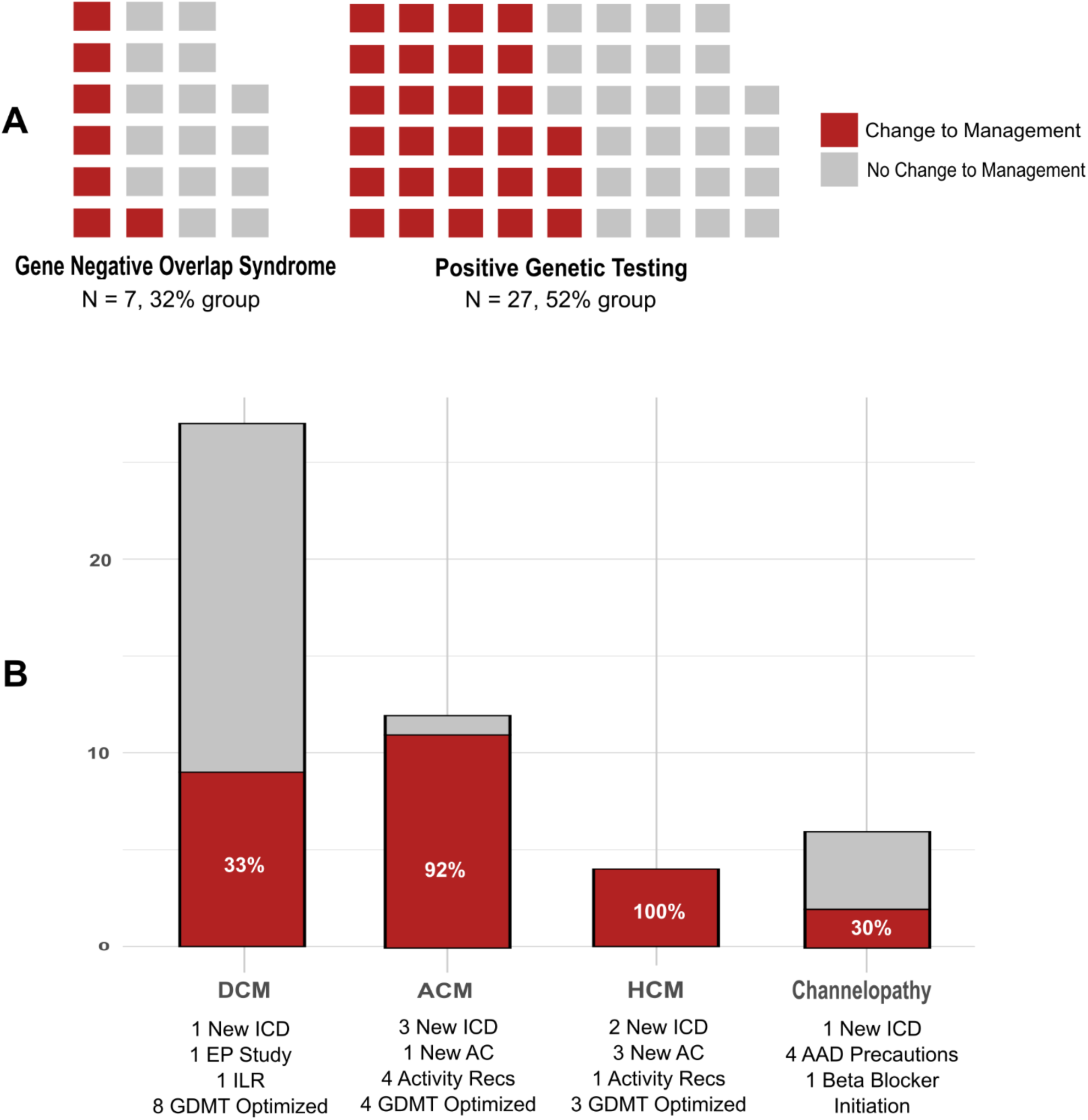
Management changes resulting from AF Precision Medicine Clinic Evaluation. **Panel A** demonstrates the proportion of participants with a change in management stratified by gene-elusive AF overlap syndrome (left figure) and those with positive genetic testing (right figure). **Panel B** shows the proportion of participants who had ≥1 change in their clinical management and summarizes the type of changes below.

## DISCUSSION

We present here our approach to clinical and genetic testing, interpretation of results, and clinical decision making in a dedicated AF precision medicine clinic. The 20% yield of positive genetic testing using a comprehensive CM/arrhythmia gene panel was twice the rate previously reported in unselected retrospective AF cohorts.(10,25,26) Another ∼10% of patients were found to have a gene elusive AF overlap syndrome, and an additional 8% of patients had a suspicious VUS. Taken together, a genetic diagnosis was made in ∼30% of patients, expected to increase with potential reclassification of suspicious VUS. The strongest clinical predictor of positive genetic testing was a history of cardiomyopathy, whereas the strongest predictor of negative genetic testing was the presence of OSA. Among phenotyping data obtained from electrocardiography and cardiac imaging, the strongest predictors of positive genetic testing were ventricular fibrosis or scar on cardiac MRI (abnormal T1 or LGE) and conduction disease (RBBB/LBBB/IVCD). A normal EKG at initial evaluation was the strongest predictor of negative genetic testing from phenotyping data. Those patients found to have positive genetic testing are susceptible to a genetic overlap syndrome, however only a proportion will develop it, while some will have isolated AF as the only clinical manifestation of genetic disease. From longitudinal retrospective data, we previously reported that the penetrance of DCM for patients with AF and pathogenic *TTN* variants was ∼50%.(18) Here, we found the penetrance varied according to the AF genetic subgroup and for most it was ∼40 to 50% (DCM genes, HCM genes, and channelopathy genes). However, the penetrance for ACM genes was higher at ∼90%. The results of genetic testing changed clinical management for 52% of patients with positive genetic testing, highlighted by 7 patients in whom a new ICD was placed and 18 that had optimization or initiation of guideline directed medical therapy.

### Yield of Genetic Testing

We and others have previously reported the yield of positive genetic testing in patients with early-onset AF was ∼10% in a predominately White cohort and slightly less in an ethnically diverse cohort(10,27); however, we report here that it was twice as high (20%) in patients referred to our clinic. Two major differences may account for the higher rate of positive genetic testing. First, younger age at AF diagnosis is associated with a higher likelihood of positive genetic testing and the median age of AF diagnosis for patients referred to our clinic was 39 years of age compared to 50 years of age for our prior retrospective study.(10) Second, participants in this study were specifically referred to our clinic for genetic evaluation by clinicians who may have had an elevated level of suspicion for a genetic etiology. The clinical acumen of referring providers enriched our prospective cohort compared to the relatively unselected retrospective cohorts used in prior reports. Our results are consistent with another case series with median age at AF diagnosis 27 years old prospectively referred to a genetics clinic that found 24% had positive genetic testing.(26) As healthcare systems and providers consider incorporating new recommendations for genetic testing in patients with early-onset AF(11,12), a yield of positive genetic testing of 20% for AF is comparable to that of other cardiac phenotypes for which genetic testing is already routinely used, such as DCM and Brugada Syndrome.(28,29)

### Gene Elusive AF Overlap Syndromes

Patients who are clinically diagnosed with an AF-overlap syndrome, but have negative genetic testing are termed gene elusive. The percentage of patients with gene elusive DCM is estimated to be ∼50-80%, ACM is ∼40-50%, HCM is ∼40%, LQTS is ∼15-30%, and Brugada Syndrome is ∼80%. The rate of gene elusive syndromes are known to be even higher in patients from non-European genetic ancestry.(12,21–24) In our study, 22 participants (9%) were found to have a gene elusive AF overlap syndrome, which included 5 (23%) from a non-European genetic ancestry.

Among the gene elusive participants, 4 (18%) had a suspicious VUS and 7 (32%) had a positive family history suspicious for an inherited cardiomyopathy/arrhythmia syndrome. Taken together, 50% of the gene elusive participants had no suspicious genetic variant or family history to suggest an AF genetic overlap syndrome, supporting the added benefit of clinical testing rather than relying on genetic testing alone. Furthermore, 4 of the gene elusive participants had abnormalities on cardiac MRI that were not appreciated on TTE, which suggests the need for additional research to identify patients that would benefit most from cardiac MRI given its higher cost and limited availability.

### Predictors of Positive and Negative Genetic Testing at the Time of Initial Clinical Evaluation

Using clinical information available during the initial evaluation, the strongest predictors of positive genetic testing were personal history of ventricular arrhythmias (sustained VT/VF > non-sustained VT or couplets) or ventricular cardiomyopathy. These suggest the presence of a pathogenic variant because they are manifestations of a ventricular phenotype of the cardiomyopathy genes sequenced in our panel.(14) Notably, family history of AF was not a predictor of positive genetic testing, likely because AF has a complex genetic architecture with polygenic risk and shared clinical risk factors that contribute to heritability rather than rare, large-effect sized variants.(12)

Alternatively, strong clinical risk factors for AF are expected to lower the yield of genetic testing.(14) Consistent with this idea, the strongest predictors of negative genetic testing were OSA, tall height (>99^th^ percentile), and alcohol use disorder. However, only OSA was statistically significant and even among patients with OSA, a relatively large proportion of them (14%) had positive genetic testing. The yield of genetic testing was similarly large in other special AF populations, such as tall patients (10%), those with alcohol use disorder (24%), endurance athletes (16%), thyroid disorder (21%), moderate obesity (BMI 30-39 kg/m^2^; 14%) and morbid obesity (BMI ≥ 40 kg/m^2^; 23%). This suggests that rare pathogenic variants confer a genetic susceptibility to AF that still requires a ‘second hit’ to develop AF.(12) In many cases, the second hits are common clinical risk factors and genetic testing should not be ruled out solely due to their presence.

### Predictors of Positive and Negative Genetic Testing from Electrocardiographic and Imaging Evaluation

As part of the phenotypic evaluation for genetic disease, patients underwent electrocardiography, cardiac imaging, and ambulatory monitoring. Conduction disease on a 12 lead ECG manifesting as LBBB, RBBB, or IVCD was a significant predictor of positive genetic testing as many genes encode for proteins responsible for electrical transduction that may also couple with impaired mechanical force dispersion associated with cardiomyopathy. One of the strongest predictors of negative genetic testing was a normal 12-lead ECG. Careful interpretation should be used to not overlook subtle ECG abnormalities found in early-stage genetic cardiomyopathies such as abnormal QRS voltage (low or high) and fascicular blocks.(18)

Using information from cardiac imaging, measurement of ventricular fibrosis from cardiac MRI was found to be one of the strongest predictors of positive genetic testing. Specifically, elevated T1 time on parametric mapping and ventricular LGE that both indicate the presence of ventricular fibrosis. Interestingly, the association for abnormal T1 appears stronger than LGE, consistent with research in patients with non-ischemic cardiomyopathy showing T1 mapping is superior to LGE for assessment of diffuse fibrosis.(30) In this clinical population, 71% (34 out of 48) of participants with positive genetic testing harbored a cardiomyopathy variant that may benefit from early detection of adverse remodeling through T1 mapping rather than LGE alone. Measurement of ventricular dilation or systolic dysfunction did not predict positive genetic testing. Although not significant, RV dilation and concentric LVH were predictors of negative genetic testing. These imaging features are consistent with diastolic dysfunction and suggest adverse atrial remodeling that predisposes to development of AF, and when coupled with shared metabolic risk factors may be more suggestive of negative genetic testing. From ambulatory monitoring, complex ventricular ectopy (non-sustained VT or ventricular couplets) was a non-significant predictor of positive genetic testing, with the understanding that the predictive utility of ventricular ectopy likely exists on a spectrum whereby increasing frequency and duration of ectopy confers additional risk beyond isolated couplets as has been suggested from other retrospective work.(31)

### Penetrance of the Ventricular Phenotype for AF Genetic Overlap Syndromes

For participants with positive genetic testing, the median time from AF diagnosis to evaluation in clinic was ∼4 years, but the absolute range was 1 week up to 43 years. This variability may affect penetrance estimates because prior studies suggest the time course, or natural history, of developing the ventricular phenotype for specific cardiomyopathy genes may significantly differ. For example, HCM may present earlier in genotype-positive patients (*MYH7*, *MYBPC3*) than DCM does in patients with pathogenic *TTN* variants. This is supported by a study that found 47% of family members with a pathogenic HCM variant were diagnosed with HCM upon initial clinical evaluation and only 3% of genotype-positive family members developed incident HCM over long term follow-up.(32) Conversely, P/LP variants associated with DCM may present later in life, as other studies have found pathogenic *TTN* variants contribute to a significant proportion of late-onset DCM (>60 years of age).(33,34)

We found the penetrance of a ventricular phenotype was 54% (13 out of 24) at the time of initial evaluation for AF patients with a pathogenic *TTN* variant. This is consistent with prior studies from our group and others.(18,35) In our prior work, 57 participants with early-onset AF and pathogenic *TTN* variants were identified with retrospective research sequencing. Over a median follow-up of 8 years (Q1, Q3: 5, 14 years), 47% developed ventricular cardiomyopathy in the form of LV systolic dysfunction and/or sustained ventricular arrhythmias. The penetrance at initial evaluation is relatively similar in these smaller HCM-gene and channelopathy-gene subgroups, 40% (2 out of 6) and 50% (2 out of 4), respectively. However, the penetrance of the ACM-gene subgroup was higher at ∼90% (8 out of 9), consistent with other studies that have found the penetrance of ACM genes, predominantly *LMNA, FLNC,* and *DSP,* are higher than the penetrance of *TTN* for DCM.(33,36)

### Changes to Clinical Management for Patients with Positive Genetic Testing

We report the clinical management that resulted from positive genetic testing by facilitating the diagnosis of an AF genetic overlap syndrome. Overall, 52% of patients with positive genetic testing had a change in their clinical management.

One of the most consequential decisions in caring for patients with genetic cardiomyopathy and arrhythmia syndromes is whether to implant an ICD. In patients with early-onset AF who undergo genetic testing, positive results can introduce special indications for ICD and pacemaker placement. First, ICD placement is considered reasonable in genetic cardiomyopathies associated with *LMNA*, *FLNC*, *PLN* and desmosomal genes if the LVEF is <45%.(37–39) One patient in our study underwent new ICD implantation due to this indication. Second, for patients diagnosed with ACM who experience syncope associated with ventricular arrhythmias, an ICD is reasonable.(37–39) One patient in our study underwent new ICD implantation due to this indication. Third, for patients diagnosed with HCM, special primary prevention ICD indications for HCM apply.(39,40) Two patients in our study were given a new diagnosis of HCM and underwent new ICD implant. Finally, for patients with an indication for permanent pacing who have a pathogenic variant associated with a high-risk for ventricular arrhythmias, an ICD with pacing capabilities is reasonable.(37) This indication applied to one patient in our study who underwent a new ICD implant and 2 patients who underwent an upgrade from a pacemaker to ICD. Special indications exist for other genetic syndromes but were not encountered for our current study.

For most patients with AF, anticoagulation for stroke prophylaxis is guided by the CHADS2-Vasc score or other similar risk estimators. However, special recommendations exist to anticoagulate patients with HCM or ACM regardless of baseline embolic stroke risk predicted by conventional risk calculators.(37,40) In our study, 7 participants with a CHADS2-Vasc score of ≤1 were given a new diagnoses of HCM or ACM and started on anticoagulation for stroke prophylaxis.

It is currently unknown how long to continue GDMT in patients who have depressed LV function in the setting of AF that recovers with successful rhythm control therapy (e.g., arrhythmia-induced or tachycardia-induced cardiomyopathy). We have previously shown that for patients with AF and pathogenic *TTN* variants who developed any degree of LV systolic dysfunction, 80% had symptomatic heart failure at the end of follow-up, among which approximately 1/3^rd^ had NYHA Class III/IV heart failure or had undergone cardiac transplant.(18) Patients with *TTN* (+) DCM are known to respond well to GDMT, thus our practice was to proactively optimize GDMT for patients with pathogenic *TTN* or other cardiomyopathy variants who had any history of LV systolic dysfunction.(41) Some patients were already on optimal GDMT at the time of evaluation, but 7 patients were not and following shared decision-making, all of them were started on beta-blockers and/or ACEi/ARB’s to prevent recurrence or progression of LV systolic dysfunction.

The association between exercise and development of AF demonstrates a U-shaped curve. On the low end, obesity and its related conditions (OSA, diabetes, hypertension) are major contributors to AF risk and treatment with weight-loss and exercise has been shown to reduce symptomatic AF and its progression.(42,43) On the other end of the spectrum, intense physical training is a risk factor for the development of AF and ongoing multicenter trials are studying specific recommendations for exercise in endurance athletes with AF.(44) In patients with genetic cardiomyopathy and arrhythmia syndromes, guidance regarding exercise restriction is becoming more lenient as emerging data support the safety of mild/moderate exercise for specific syndromes (e.g., non-obstructive HCM, Long QT Syndrome).(45,46) However, concern remains that intense, endurance exercise can promote development or progression of disease, and participation should only be considered in consultation with a provider specialized in these conditions.(47,48) The strongest data support exercise restriction in patients with pathogenic desmosomal variants with ARVC, but these results have been generalized to also include LV and biventricular forms of ACM.(37) The specific recommendation is to encourage patients with ACM, or a pathogenic ACM variant, to limit physical activity to ≤6 metabolic equivalents.(37) This affected four participants with ACM in our study.

Patients with Long QT Syndrome or Brugada Syndrome, or pathogenic variants in genes associated with either syndrome, are recommended to avoid medications that prolong QT or block the cardiac potassium or sodium channels, respectively.(39,47) We recommended avoidance of Class I antiarrhythmics (e.g., flecainide, propafenone), which block the cardiac sodium channel, in three patients with pathogenic *SCN5A* variants. Avoidance of Class III antiarrhythmics (e.g., sotalol, dofetilide), which block cardiac potassium channels, was also recommended in one participant with a pathogenic *KCNQ1* variant.

### Limitations

Approximately 90% of the study participants were White/non-Hispanic, limiting the generalizability of these findings to more diverse populations. The sample size for many subgroup analyses was relatively small, which may affect the accuracy of the descriptive statistics. Polygenic risk is a major contributor to genetic susceptibility to AF, which was not defined in this study. There is considerable phenotypic and genetic overlap between DCM and ACM and the definitions for each are evolving and differ between different professional societies.

## CONCLUSION

Here, we describe our current framework for evaluation of patients presenting with early onset AF in a dedicated AF precision medicine clinic. We demonstrate that features from the patient’s history can raise suspicion for genetic disease, and that the presence of clinical risk factors for AF does not exclude a genetic diagnosis. Genetic evaluation and detailed phenotyping have important diagnostic and clinical management implications for patients with early onset atrial fibrillation.

## SUPPLEMENTARY DATA

Section S.1: Supplementary Methods

Supplemental Table 1: Complete list of genes grouped according to AF genetic subgroups

Supplemental Table 2: Diagnostic criteria and management considerations for Inherited Syndromes

Supplemental Figure 1: AFPMC Best Practice Alert through Electronic Medical Record

Supplemental Table 3: Details of genetic testing and full summary of results

Supplemental Table 4: Suspicious variants of undetermined significance (VUS)

Supplemental Table 5: Details of clinical testing

Supplemental Table 6: Details of phenotype penetrance and changes to management

## ACKNOWLEDGEMENTS

This study was also made possible by support to our genetic arrhythmia program by the Shah Foundation and Cohen Foundation. We thank the cardiology, genetics and electrophysiology clinical staff for their tireless support of our program in caring for these patients.

## DATA AVAILABILITY STATEMENT

The data underlying this article cannot be shared publicly for the privacy of patients that participated in the registry. The data will be shared on reasonable request to the corresponding author.

## REFERENCES

1. Ohlrogge AH, Brederecke J, Schnabel RB. Global Burden of Atrial Fibrillation and Flutter by National Income: Results From the Global Burden of Disease 2019 Database. J Am Heart Assoc. 2023 Sep 5;12(17):e030438.

2. Laitinen-Forsblom PJ, Mäkynen P, Mäkynen H, Yli-Mäyry S, Virtanen V, Kontula K, et al. SCN5A mutation associated with cardiac conduction defect and atrial arrhythmias. J Cardiovasc Electrophysiol. 2006 May;17(5):480–5.

3. Chen YH, Xu SJ, Bendahhou S, Wang XL, Wang Y, Xu WY, et al. KCNQ1 gain-of-function mutation in familial atrial fibrillation. Science. 2003 Jan 10;299(5604):251–4.

4. Pan H, Richards AA, Zhu X, Joglar JA, Yin HL, Garg V. A novel mutation in LAMIN A/C is associated with isolated early-onset atrial fibrillation and progressive atrioventricular block followed by cardiomyopathy and sudden cardiac death. Heart Rhythm. 2009 May;6(5):707–10.

5. Ackerman MJ, Priori SG, Willems S, Berul C, Brugada R, Calkins H, et al. HRS/EHRA expert consensus statement on the state of genetic testing for the channelopathies and cardiomyopathies: this document was developed as a partnership between the Heart Rhythm Society (HRS) and the European Heart Rhythm Association (EHRA). Eur Eur Pacing Arrhythm Card Electrophysiol J Work Groups Card Pacing Arrhythm Card Cell Electrophysiol Eur Soc Cardiol. 2011 Aug;13(8):1077–109.

6. Choi SH, Weng LC, Roselli C, Lin H, Haggerty CM, Shoemaker MB, et al. Association Between Titin Loss-of-Function Variants and Early-Onset Atrial Fibrillation. JAMA. 2018 Dec 11;320(22):2354–64.

7. Ahlberg G, Refsgaard L, Lundegaard PR, Andreasen L, Ranthe MF, Linscheid N, et al. Rare truncating variants in the sarcomeric protein titin associate with familial and early-onset atrial fibrillation. Nat Commun. 2018 Oct 17;9(1):4316.

8. Tomaselli GF, Roy-Puckelwartz MJ. Early-Onset Atrial Fibrillation and Heritable Heart Disease-To Test or Not to Test? JAMA Cardiol. 2021 Dec 1;6(12):1359–61.

9. Shoemaker MB, Shah RL, Roden DM, Perez MV. How Will Genetics Inform the Clinical Care of Atrial Fibrillation? Circ Res. 2020 Jun 19;127(1):111–27.

10. Yoneda ZT, Anderson KC, Quintana JA, O’Neill MJ, Sims RA, Glazer AM, et al. Early-Onset Atrial Fibrillation and the Prevalence of Rare Variants in Cardiomyopathy and Arrhythmia Genes. JAMA Cardiol. 2021 Dec 1;6(12):1371–9.

11. Joglar JA, Chung MK, Armbruster AL, Benjamin EJ, Chyou JY, Cronin EM, et al. 2023 ACC/AHA/ACCP/HRS Guideline for the Diagnosis and Management of Atrial Fibrillation: A Report of the American College of Cardiology/American Heart Association Joint Committee on Clinical Practice Guidelines. Circulation. 2024 Jan 2;149(1):e1–156.

12. Wilde AAM, Semsarian C, Márquez MF, Shamloo AS, Ackerman MJ, Ashley EA, et al. European Heart Rhythm Association (EHRA)/Heart Rhythm Society (HRS)/Asia Pacific Heart Rhythm Society (APHRS)/Latin American Heart Rhythm Society (LAHRS) Expert Consensus Statement on the state of genetic testing for cardiac diseases. Eur Eur Pacing Arrhythm Card Electrophysiol J Work Groups Card Pacing Arrhythm Card Cell Electrophysiol Eur Soc Cardiol. 2022 Sep 1;24(8):1307–67.

13. Yoneda ZT, Anderson KC, Ye F, Quintana JA, O’Neill MJ, Sims RA, et al. Mortality Among Patients With Early-Onset Atrial Fibrillation and Rare Variants in Cardiomyopathy and Arrhythmia Genes. JAMA Cardiol. 2022 Jul 1;7(7):733–41.

14. Kany S, Jurgens SJ, Rämö JT, Christophersen IE, Rienstra M, Chung MK, et al. Genetic testing in early-onset atrial fibrillation. Eur Heart J. 2024 Sep 7;45(34):3111–23.

15. McNally EM, Khan SS. Genetic Testing for Early-Onset Atrial Fibrillation-Is It Time to Personalize Care? JAMA Cardiol. 2022 Jul 1;7(7):669–71.

16. Richards S, Aziz N, Bale S, Bick D, Das S, Gastier-Foster J, et al. Standards and guidelines for the interpretation of sequence variants: a joint consensus recommendation of the American College of Medical Genetics and Genomics and the Association for Molecular Pathology. Genet Med Off J Am Coll Med Genet. 2015 May;17(5):405–24.

17. Roberts AM, Ware JS, Herman DS, Schafer S, Baksi J, Bick AG, et al. Integrated allelic, transcriptional, and phenomic dissection of the cardiac effects of titin truncations in health and disease. Sci Transl Med. 2015 Jan 14;7(270):270ra6.

18. Virk ZM, El-Harasis MA, Yoneda ZT, Anderson KC, Sun L, Quintana JA, et al. Clinical Characteristics and Outcomes in Patients With Atrial Fibrillation and Pathogenic TTN Variants. JACC Clin Electrophysiol. 2024 Sep 27;S2405-500X(24)00765-5.

19. Puckelwartz MJ, Pesce LL, Hernandez EJ, Webster G, Dellefave-Castillo LM, Russell MW, et al. The impact of damaging epilepsy and cardiac genetic variant burden in sudden death in the young. Genome Med. 2024 Jan 16;16(1):13.

20. Arbelo E, Protonotarios A, Gimeno JR, Arbustini E, Barriales-Villa R, Basso C, et al. 2023 ESC Guidelines for the management of cardiomyopathies. Eur Heart J. 2023 Oct 1;44(37):3503–626.

21. Jordan E, Kinnamon DD, Haas GJ, Hofmeyer M, Kransdorf E, Ewald GA, et al. Genetic Architecture of Dilated Cardiomyopathy in Individuals of African and European Ancestry. JAMA. 2023 Aug 1;330(5):432–41.

22. Fatumo S, Chikowore T, Choudhury A, Ayub M, Martin AR, Kuchenbaecker K. A roadmap to increase diversity in genomic studies. Nat Med. 2022 Feb;28(2):243–50.

23. Rosamilia MB, Markunas AM, Kishnani PS, Landstrom AP. Underrepresentation of Diverse Ancestries Drives Uncertainty in Genetic Variants Found in Cardiomyopathy-Associated Genes. JACC Adv. 2024 Feb;3(2):100767.

24. Landry LG, Ali N, Williams DR, Rehm HL, Bonham VL. Lack Of Diversity In Genomic Databases Is A Barrier To Translating Precision Medicine Research Into Practice. Health Aff Proj Hope. 2018 May;37(5):780–5.

25. Chalazan B, Mol D, Darbar FA, Ornelas-Loredo A, Al-Azzam B, Chen Y, et al. Association of Rare Genetic Variants and Early-Onset Atrial Fibrillation in Ethnic Minority Individuals. JAMA Cardiol. 2021 Jul 1;6(7):811–9.

26. Goodyer WR, Dunn K, Caleshu C, Jackson M, Wylie J, Moscarello T, et al. Broad Genetic Testing in a Clinical Setting Uncovers a High Prevalence of Titin Loss-of-Function Variants in Very Early Onset Atrial Fibrillation. Circ Genomic Precis Med. 2019 Nov;12(11):e002713.

27. Chalazan B, Mol D, Darbar FA, Ornelas-Loredo A, Al-Azzam B, Chen Y, et al. Association of Rare Genetic Variants and Early-Onset Atrial Fibrillation in Ethnic Minority Individuals. JAMA Cardiol. 2021 Jul 1;6(7):811–9.

28. Verdonschot JAJ, Hazebroek MR, Krapels IPC, Henkens MTHM, Raafs A, Wang P, et al. Implications of Genetic Testing in Dilated Cardiomyopathy. Circ Genomic Precis Med. 2020 Oct;13(5):476–87.

29. Pannone L, Bisignani A, Osei R, Gauthey A, Sorgente A, Monaco C, et al. Genetic Testing in Brugada Syndrome: A 30-Year Experience. Circ Arrhythm Electrophysiol. 2024 Apr;17(4):e012374.

30. Sramko M, Abdel-Kafi S, Wijnmaalen AP, Tao Q, van der Geest RJ, Lamb HJ, et al. Head-to-Head Comparison of T1 Mapping and Electroanatomical Voltage Mapping in Patients With Ventricular Arrhythmias. JACC Clin Electrophysiol. 2023 Jun;9(6):740–8.

31. Setti M, Merlo M, Gigli M, Munaretto L, Paldino A, Stolfo D, et al. Role of arrhythmic phenotype in prognostic stratification and management of dilated cardiomyopathy. Eur J Heart Fail. 2024;26(3):581–9.

32. Schoonvelde SAC, Alexandridis GM, Price LB, Schinkel AFL, Hirsch A, Zwetsloot PP, et al. Family screening for hypertrophic cardiomyopathy: Initial cardiologic assessment, and long-term follow-up of genotype-positive phenotype-negative individuals. Int J Cardiol. 2024 Dec 31;422:132951.

33. Cabrera-Romero E, Ochoa JP, Barriales-Villa R, Bermúdez-Jiménez FJ, Climent-Payá V, Zorio E, et al. Penetrance of Dilated Cardiomyopathy in Genotype-Positive Relatives. J Am Coll Cardiol. 2024 Apr 30;83(17):1640–51.

34. Cannatà A, Merlo M, Dal Ferro M, Barbati G, Manca P, Paldino A, et al. Association of Titin Variations With Late-Onset Dilated Cardiomyopathy. JAMA Cardiol. 2022 Apr 1;7(4):371–7.

35. Schiabor Barrett KM, Cirulli ET, Bolze A, Rowan C, Elhanan G, Grzymski JJ, et al. Cardiomyopathy prevalence exceeds 30% in individuals with TTN variants and early atrial fibrillation. Genet Med Off J Am Coll Med Genet. 2023 Apr;25(4):100012.

36. Kumar S, Baldinger SH, Gandjbakhch E, Maury P, Sellal JM, Androulakis AFA, et al. Long-Term Arrhythmic and Nonarrhythmic Outcomes of Lamin A/C Mutation Carriers. J Am Coll Cardiol. 2016 Nov 29;68(21):2299–307.

37. Towbin JA, McKenna WJ, Abrams DJ, Ackerman MJ, Calkins H, Darrieux FCC, et al. 2019 HRS expert consensus statement on evaluation, risk stratification, and management of arrhythmogenic cardiomyopathy. Heart Rhythm. 2019 Nov;16(11):e301–72.

38. Heidenreich PA, Bozkurt B, Aguilar D, Allen LA, Byun JJ, Colvin MM, et al. 2022 AHA/ACC/HFSA Guideline for the Management of Heart Failure: A Report of the American College of Cardiology/American Heart Association Joint Committee on Clinical Practice Guidelines. Circulation. 2022 May 3;145(18):e895–1032.

39. Al-Khatib SM, Stevenson WG, Ackerman MJ, Bryant WJ, Callans DJ, Curtis AB, et al. 2017 AHA/ACC/HRS Guideline for Management of Patients With Ventricular Arrhythmias and the Prevention of Sudden Cardiac Death: Executive Summary: A Report of the American College of Cardiology/American Heart Association Task Force on Clinical Practice Guidelines and the Heart Rhythm Society. Circulation. 2018 Sep 25;138(13):e210–71.

40. Ommen SR, Ho CY, Asif IM, Balaji S, Burke MA, Day SM, et al. 2024 AHA/ACC/AMSSM/HRS/PACES/SCMR Guideline for the Management of Hypertrophic Cardiomyopathy: A Report of the American Heart Association/American College of Cardiology Joint Committee on Clinical Practice Guidelines. Circulation. 2024 Jun 4;149(23):e1239–311.

41. Akhtar MM, Lorenzini M, Cicerchia M, Ochoa JP, Hey TM, Sabater Molina M, et al. Clinical Phenotypes and Prognosis of Dilated Cardiomyopathy Caused by Truncating Variants in the *TTN* Gene. Circ Heart Fail [Internet]. 2020 Oct [cited 2024 Apr 21];13(10). Available from: https://www.ahajournals.org/doi/10.1161/CIRCHEARTFAILURE.119.006832

42. Abed HS, Wittert GA, Leong DP, Shirazi MG, Bahrami B, Middeldorp ME, et al. Effect of weight reduction and cardiometabolic risk factor management on symptom burden and severity in patients with atrial fibrillation: a randomized clinical trial. JAMA. 2013 Nov 20;310(19):2050–60.

43. Wang TJ, Parise H, Levy D, D’Agostino RB, Wolf PA, Vasan RS, et al. Obesity and the risk of new-onset atrial fibrillation. JAMA. 2004 Nov 24;292(20):2471–7.

44. Apelland T, Janssens K, Loennechen JP, Claessen G, Sørensen E, Mitchell A, et al. Effects of training adaption in endurance athletes with atrial fibrillation: protocol for a multicentre randomised controlled trial. BMJ Open Sport Exerc Med. 2023;9(2):e001541.

45. Tobert KE, Bos JM, Garmany R, Ackerman MJ. Return-to-Play for Athletes With Long QT Syndrome or Genetic Heart Diseases Predisposing to Sudden Death. J Am Coll Cardiol. 2021 Aug 10;78(6):594– 604.

46. Gudmundsdottir HL, Axelsson Raja A, Rossing K, Rasmusen H, Snoer M, Andersen LJ, et al. Exercise Training in Patients With Hypertrophic Cardiomyopathy Without Left Ventricular Outflow Tract Obstruction: A Randomized Clinical Trial. Circulation. 2025 Jan 14;151(2):132–44.

47. Priori SG, Wilde AA, Horie M, Cho Y, Behr ER, Berul C, et al. HRS/EHRA/APHRS expert consensus statement on the diagnosis and management of patients with inherited primary arrhythmia syndromes: document endorsed by HRS, EHRA, and APHRS in May 2013 and by ACCF, AHA, PACES, and AEPC in June 2013. Heart Rhythm. 2013 Dec;10(12):1932–63.

48. Dagradi F, Spazzolini C, Castelletti S, Pedrazzini M, Kotta MC, Crotti L, et al. Exercise Training-Induced Repolarization Abnormalities Masquerading as Congenital Long QT Syndrome. Circulation. 2020 Dec 22;142(25):2405–15.

49. Harris PA, Taylor R, Thielke R, Payne J, Gonzalez N, Conde JG. Research electronic data capture (REDCap)--a metadata-driven methodology and workflow process for providing translational research informatics support. J Biomed Inform. 2009 Apr;42(2):377–81.

50. Heidenreich PA, Bozkurt B, Aguilar D, Allen LA, Byun JJ, Colvin MM, et al. 2022 AHA/ACC/HFSA Guideline for the Management of Heart Failure. J Am Coll Cardiol. 2022 May;79(17):e263–421.

51. Marcus FI, McKenna WJ, Sherrill D, Basso C, Bauce B, Bluemke DA, et al. Diagnosis of arrhythmogenic right ventricular cardiomyopathy/dysplasia: Proposed Modification of the Task Force Criteria. Eur Heart J. 2010 Apr 1;31(7):806–14.

52. Kowdley KV, Brown KE, Ahn J, Sundaram V. ACG Clinical Guideline: Hereditary Hemochromatosis. Off J Am Coll Gastroenterol ACG. 2019 Aug;114(8):1202.

53. Contributors and Hemochromatosis International Taskforce, Adams P, Altes A, Brissot P, Butzeck B, Cabantchik I, et al. Therapeutic recommendations in HFE hemochromatosis for p.Cys282Tyr (C282Y/C282Y) homozygous genotype. Hepatol Int. 2018 Mar;12(2):83–6.

54. Kittleson MM, Ruberg FL, Ambardekar AV, Brannagan TH, Cheng RK, Clarke JO, et al. 2023 ACC Expert Consensus Decision Pathway on Comprehensive Multidisciplinary Care for the Patient With Cardiac Amyloidosis: A Report of the American College of Cardiology Solution Set Oversight Committee. J Am Coll Cardiol. 2023 Mar 21;81(11):1076–126.

