## Supplemental Data for "The Therapeutic Impact of Genetic Evaluation in an Atrial Fibrillation Precision Medicine Clinic"

**Atrial Fibrillation Precision Medicine Clinic**

**SUPPLEMENTARY DATA**

Section S.1: Supplementary Methods

Supplemental Table 1: Complete list of genes grouped according to AF genetic subgroups

Supplemental Table 2: Diagnostic criteria and management considerations for Inherited Syndromes

Supplemental Figure 1: AFPMC Best Practice Alert through Electronic Medical Record

Supplemental Table 3: Details of genetic testing and full summary of results

Supplemental Table 4: Suspicious variants of undetermined significance (VUS)

Supplemental Table 5: Details of clinical testing

Supplemental Table 6: Details of phenotype penetrance and changes to management

**Supplementary Methods S.1**

Referral to the AF Precision Medicine Clinic

Referral to the clinic was placed by primary providers with a clinical suspicion for a genetic etiology of their patient’s atrial fibrillation and associated cardiac comorbid conditions. A best practice alert through the electronic medical record was also implemented for young patients with atrial fibrillation that did not have significant predisposing factors (congenital heart disease, severe valvular disease, etc.) to facilitate referral of appropriate patients that may benefit from genetic evaluation (**Supplemental Figure 1**). Patients in the clinic were prospectively enrolled in an IRB-approved registry (IRB #201666, NCT05190679). Participant data were recorded in a password-protected database designed for clinical research (REDCap).(49) Some family members underwent targeted genotyping limited to only the familial variant. Depending on the results of the standard clinical evaluation and genetic testing, additional diagnostic testing with an exercise tolerance test, extended ECG monitoring, or a procainamide challenge was performed. Patients were seen for a 3-month follow-up appointment either in-person or by telemedicine to review the results of their tests and develop a longitudinal care plan.

Variant Interpretation

The clinical genetic testing laboratories and investigators used the ACMG/AMP (American College of Medical Genetics and Association for Molecular Pathology) Standards and Guidelines for variant interpretation.(16) The ACMG/AMP criteria designate a variant as pathogenic (P), likely-pathogenic (LP), a variant of undetermined significance (VUS), likely-benign (LB), or benign (B). If any variants were officially reclassified by the clinical genetic testing laboratory, the current classification as of 2/20/2025 was used for this analysis.

“Positive” genetic testing was defined by any of the following conditions:

- One or more P/LP variants in a gene with autosomal dominant (AD) inheritance, or
- Two P/LP variants in a gene with autosomal recessive (AR) inheritance, or
- One P/LP variant in hemizygous men or homozygous women for genes with X-linked inheritance.

AF Genetic Subgroups

Genes were classified according to the predominant gene-phenotype association here termed “AF genetic subgroups”. This a challenging clinical issue, as there is often overlapping genetic architecture of distinct phenotypes (eg ACM/NDLVC genes may have LV dilation as with DCM), and also different variants within the same gene can cause different phenotypes (eg *MYH7* is both a definitive evidence gene for DCM and HCM.) Categorized of “AF genetic subgroups” was done by gene-level predominant phenotypic association to facilitate patient management according to genetic susceptibility for syndromes with established guidelines that inform clinical management. Genes associated with syndromes that are less common in our study population and genes with only limited, preliminary, refuted, or disputed evidence for their gene-disease association according to ClinGen were not assigned to an AF genetic subgroup.

**SUPPLEMENTAL TABLE 1: Complete list of genes (N=175) for comprehensive cardiac genetic testing panels.** Individual genes tested varied by vendor. This table lists the most comprehensive gene panel utilized by the Vanderbilt Clinical Genetics Laboratory.

| *A2ML1*  *ABCC9*  *ACADVL*  *ACTC1*  *ACTN2*  *AGL*  *AKAP9*  *ALMS1*  *ALPK3*  *ANK2*  *ANKRD1*  *BAG3*  *BRAF*  *CACNA1C*  *CACNA2D1*  *CACNB2*  *CALM1*  *CALM2*  *CALM3*  *CALR3*  *CASQ2*  *CAV3*  *CAVIN4*  *CBL*  *CHRM2*  *CPT2*  *CRYAB*  *CSRP3*  *CTF1*  *CTNNA3*  *DEPDC5*  *DES*  *DMD*  *DNAJC19*  *DOLK* | *DSC2*  *DSG2*  *DSP*  *DTNA*  *ELAC2*  *EMD*  *EYA4*  *FHL1*  *FHL2*  *FKRP*  *FKTN*  *FLNC*  *GAA*  *GATA4*  *GATA5*  *GATA6*  *GATAD1*  *GJA5*  *GLA*  *GNB5*  *GPD1L*  *HCN4*  *HFE*  *HRAS*  *ILK*  *JPH2*  *JUP*  *KCNA1*  *KCNA5*  *KCND3*  *KCNE1*  *KCNE2*  *KCNE3*  *KCNE5*  *KCNH2* | *KCNJ2*  *KCNJ5*  *KCNJ8*  *KCNK3*  *KCNQ1*  *KCNQ2*  *KCNQ3*  *KCNT1*  *KRAS*  *LAMA4*  *LAMP2*  *LDB3*  *LMNA*  *LRRC10*  *MAP2K1*  *MAP2K2*  *MED12*  *MIB1*  *MTND1*  *MTND5*  *MTND6*  *MTO1*  *MTTD*  *MTTG*  *MTTH*  *MTTI*  *MTTK*  *MTTL1*  *MTTL2*  *MTTM*  *MTTQ*  *MTTS1*  *MTTS2*  *MYBPC3*  *MYH6* | *MYH7*  *MYL2*  *MYL3*  *MYL4*  *MYLK2*  *MYOM1*  *MYOZ2*  *MYPN*  *NEBL*  *NEXN*  *NF1*  *NKX2-5*  *NPPA*  *NRAS*  *PCDH19*  *PDLIM3*  *PKP2*  *PLEKHM2*  *PLN*  *PPA2*  *PRDM16*  *PRKAG2*  *PRRT2*  *PTPN11*  *RAF1*  *RANGRF*  *RASA1*  *RBM20*  *RIT1*  *RRAS*  *RYR2*  *SCN10A*  *SCN1A*  *SCN1B*  *SCN2B* | *SCN3B*  *SCN4B*  *SCN5A*  *SCN8A*  *SCN9A*  *SDHA*  *SGCD*  *SHOC2*  *SLC22A5*  *SLC2A1*  *SLMAP*  *SNTA1*  *SOS1*  *SOS2*  *SPRED1*  *TAZ*  *TBX20*  *TBX5*  *TCAP*  *TECRL*  *TGFB3*  *TMEM43*  *TMEM70*  *TMPO*  *TNNC1*  *TNNI3*  *TNNT2*  *TOR1AIP1*  *TPM1*  *TRDN*  *TRPM4*  *TTN*  *TTR*  *TXNRD2*  *VCL* |
| --- | --- | --- | --- | --- |

**SUPPLEMENTAL TABLE 2: Diagnostic criteria and management considerations used for inherited syndromes are based on international guidelines and task force recommendations. Note, Arrhythmogenic Cardiomyopathy (ACM) is used to describe both Non-Dilated LV Cardiomyopathy (NDLVC) and Arrhythmogenic Right Ventricular Cardiomyopathy (ARVC) phenotypes.**

| ***Syndrome*** | ***Diagnosis*** | ***Management Considerations*** | ***Citations*** |
| --- | --- | --- | --- |
| **Dilated Cardiomyopathy (DCM)** | LV dilation with impaired LV function (LVEF <50%) | HFrEF GDMT  SCD Risk Stratification  Stroke Risk Reduction | 2023 ESC (20)  2022 AHA/ACC/HFSA (50) |
| **Non-dilated Left Ventricular Cardiomyopathy**  **(NDLVC)** | Impaired LV systolic function without LV dilation, or LV fibrosis by DE-CMR | HFrEF GDMT  SCD Risk Stratification  Stroke Risk Reduction | 2023 ESC (20)  2022 AHA/ACC/HFSA (50) |
| **Hypertrophic Cardiomyopathy**  **(HCM)** | Increased LV wall thickness (>1.5 cm men, >1.3 cm women, >1.3 cm relative of genotype positive family member) | SCD Risk Stratification  Stroke Risk Reduction  LVOT Obstruction | 2024 AHA/ACC/  AMSSM/HRS/PACES/  SCMR (40) |
| **Arrhythmogenic Right Ventricular Cardiomyopathy (ARVC)** | *Definite:* 2 major, or 2 major + 1 minor, or 4 minor  *Borderline:* 1 major + 1 minor, or 3 minor  *Possible:* 1 major, or 2 minor | SCD Risk Stratification  Exercise Restriction  Stroke Risk Reduction | 2010 Modified Taskforce Criteria (51)  2023 ESC (20) |
| **Arrhythmogenic Cardiomyopathy**  **(ACM)** | Cardiomyopathy with arrhythmia predominant phenotype. Used in this manuscript to encompass both NDLVC and ARVC. | Per NDLVC and ARVC phenotype | 2019 HRS (37) |
| **Brugada Syndrome** | Type I EKG pattern spontaneously or with sodium channel blocker challenge | SCD Risk Stratification  Drug Precautions | 2013 HRS/EHRA/APHRS (47) |
| **Long QT Syndrome** | 1. LQTS risk score >3.5 *or*  2. Pathogenic LQTS variant *or*  3. QTc >500 ms without secondary cause | SCD Risk Stratification  Drug Precautions | 2013 HRS/EHRA/APHRS (47) |
| **Catecholaminergic Polymorphic Ventricular Tachycardia (CPVT)** | 1. Exercise-induced bidirectional VT or PMVT *or*  2. Pathogenic CPVT variant with exercise induced PVCs or VT | Exercise Restriction  SCD Risk Stratification  AAD Therapies | 2013 HRS/EHRA/APHRS  (47) |
| **Progressive Cardiac Conduction Disease (PCCD)** | Unexplained progressive conduction abnormalities age <50y in the absence of skeletal myopathies | SCD Risk Stratification | 2013 HRS/EHRA/APHRS  (47) |
| **Hemochromatosis** | Serum ferritin >200 ng/mL (women) or >300 (men), *and* Tsat > 45%, *and* two pathogenic HFE variants | Therapeutic phlebotomy | 2019 ACG (52)  2018 HFE International Taskforce (53) |
| **Amyloidosis** | Histologic evidence of TTR amyloid deposition, or imaging consistent with cardiac amyloid deposition and pathogenic TTR variant | TTR-directed therapy | 2023 ACC Expert Consensus (54) |
| HFrEF= heart failure with reduced ejection fraction. GDMT=guideline directed medical therapy. SCD=sudden cardiac death. ESC=European Society of Cardiology. AHA=American Heart Association. ACC=American College of Cardiology. HFSA=Heart Failure Society of America. DE-CMR= delayed enhancement cardiac MRI. LVOT=LV outflow tract. AMSSM= American Medical Society for Sports Medicine. HRS=Heart Rhythm Society. PACES= Pediatric and Congenital Electrophysiology Society. SCMR= Society for Cardiovascular Magnetic Resonance. EHRA=European Heart Rhythm Association. APHRS=Asian Pacific Heart Rhythm Society. ACG=American College of Gastroenterology. AAD = antiarrhythmic drug. TSAT= transferrin saturation. *HFE*=homeostatic iron regulator gene. *TTR*= transthyretin gene. | | | |

**SUPPLEMENTAL FIGURE 1:** Best practice alert example for a 37-year-old patient without other clear etiologies identified from automated ICD/CPT code review for referral to the Atrial Fibrillation Precision Medicine Clinic.

 
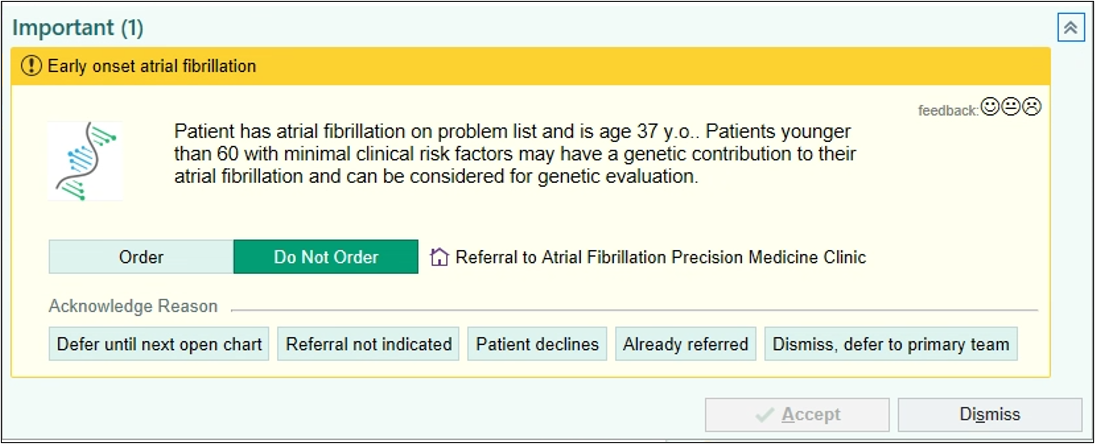

**SUPPLEMENTAL TABLE 3: Details of genetic testing and full summary of results**

|  | **Total**  **Cohort**  **(N=246*)** | **Genetic Evaluation Groups** | | |
| --- | --- | --- | --- | --- |
|  |  | **Positive**  **Genetic Testing**  **(N=52)** | **Gene Elusive AF Overlap**  **Syndrome**  **(N=22)** | **Negative**  **Genetic Evaluation**  **(N=172)** |
| Type of testing  Multi-gene CM/arrhythmia panel  Targeted genotyping | 242 (98.4%)  4 (1.6%) | 48 (92.3%)  4 (7.7%) | 22 (100%)  0 (0%) | 172 (100%)  0 (0%) |
| Genetic testing laboratory  VUMC Clinical Genetics Lab  Invitae  GeneDx  Other | 60 (24.4%)  115 (46.7%)  69 (28.0%)  2 (0.8%) | 12 (23.1%)  12 (23.1%)  16 (30.8%)  1 (1.9%) | 8 (36.4%)  8 (36.4%)  6 (27.3%)  1 (4.5%) | 40 (23.3%)  40 (23.3%)  47 (27.3%)  0 (0%) |
| Number of genes on panel | 157 [76.0, 197] | 157 [82.0, 175] | 157 [76.0, 175] | 157 [100, 197] |
| Number of variants reported | 1 [0, 6] | 2.50 [1, 5] | 1 [0, 4] | 1 [0, 6] |
| Genetic testing results  Positive  Negative, with a VUS reported  Negative, and carrier for an AR syndrome  Negative, with no rare variants reported | 52 (21.1%)  129 (52.4%)  18 (7.3%)  47 (19.1%) | 52 (100%)  0 (0%)  0 (0%)  0 (0%) | 0 (0%)  15 (68.2%)  3 (13.6%)  4 (18.2%) | 0 (0%)  114 (66.3%)  15 (8.7%)  43 (25.0%) |
| Compound or digenic heterozygote | 1 (0.4%) | 1 (1.9%) | 0 (0%) | 0 (0%) |
| VUS reported | 162 (65.9%) | 33 (63.5%) | 15 (68.2%) | 114 (66.3%) |
| “Suspicious” VUS | 36 (14.6%) | 6 (11.5%) | 4 (18.2%) | 26 (15.1%) |
| Heterozygous carrier of pathogenic AR gene | 60 (24.4%) | 13 (25.0%) | 7 (31.8%) | 40 (23.3%) |
| Tested for Myotonic Dystrophy | 31 (12.6%) | 6 (11.5%) | 3 (13.6%) | 22 (12.8%) |
| Gene for heterozygous AR carriers  *FKRP*  *GAA*  *HFE*  *SGCD*  *TRDN*  *ACADVL*  *ALMS1*  *DNAJC19*  *ELAC2*  *MYPN*  *PPA2*  *SLC22A5* | 2 (0.8%)  6 (2.4%)  42 (17.1%)  1 (0.4%)  1 (0.4%)  2 (0.8%)  1 (0.4%)  1 (0.4%)  1 (0.4%)  1 (0.4%)  1 (0.4%)  1 (0.4%) | 1 (1.9%)  1 (1.9%)  9 (17.3%)  1 (1.9%)  1 (1.9%)  0 (0%)  0 (0%)  0 (0%)  0 (0%)  0 (0%)  0 (0%)  0 (0%) | 0 (0%)  0 (0%)  7 (31.8%)  0 (0%)  0 (0%)  0 (0%)  0 (0%)  0 (0%)  0 (0%)  0 (0%)  0 (0%)  0 (0%) | 1 (0.6%)  5 (2.9%)  26 (15.1%)  0 (0%)  0 (0%)  2 (1.2%)  1 (0.6%)  1 (0.6%)  1 (0.6%)  1 (0.6%)  1 (0.6%)  1 (0.6%) |

**SUPPLEMENTAL TABLE 4: Suspicious variants of undetermined significance (VUS)**

| **Patient** | **Suspicious VUS Gene** | **P/LP Gene** | **Gene-Elusive Overlap Syndrome** | **VUS cDNA** | **VUS AA** | **Notable Family History** | **Notable Findings from Phenotypic Evaluation** |
| --- | --- | --- | --- | --- | --- | --- | --- |
| **1** | *KCNA5* | *TTN* |  | c.1327A>G | p.Ille443Val | Unremarkable | Ventricular couplets on ambulatory monitor |
| **2** | *KCNQ1* | *TTN* |  | c.1092C>G | p.Phe364Leu | Familial AF | Ventricular couplets on ambulatory monitor |
| **3** | *MYH6* | *TTN* |  | c.3382C>T | p.Arg1128Cys | HCM | LGE on CMR |
| **4** | *TTN* | *TTN* |  | c.70282 G>T | p.Val23428Leu | Familial AF, Heart Transplant | LGE, mildly reduced LV systolic function, NSVT |
| **5** | *TTN* | *TTN* |  | c.66617G>A | p.Cys22206Tyr | Familial AF | Unremarkable |
| **6** | *TTN* | *MYH7* |  | c.102790C>T | p.Leu34264Phe | Familial AF | NSVT on ambulatory monitor |
| **7** | *SCN5A* |  | DCM | c.5701G>A | p.Glu1901Lys | SUD | Mildly reduced LV systolic function |
| **8** | *TTN* |  | DCM | c.58870G>A | p.Asp19624Asn | PPM | Mildly reduced LV systolic function |
| **9** | *ACTC1* |  | ACM/NDLVC | c.309C>A | p.His103Gin | Unremarkable | LGE on CMR |
| **10** | *TRPM4* |  | ACM/NDLVC | c.2295dup | p.Arg766Alafs*193 | Unremarkable | Mildly reduced LV systolic function |
| **11** | *ABCC9* |  |  | c.1130T>C | p.Ile377Thr | Familial AF | Unremarkable |
| **12** | *ABCC9,*  *DES* |  |  | c.2408C>T, c.1023+6T>G | p.Thr803Ile, Intronic | Familial AF | NSVT on ambulatory monitor |
| **13** | *ACTC2* |  |  | c.1907 A>G | p.GLu636Gly | Familial AF, SUD | NSVT on ambulatory monitor |
| **14** | *ACTN2* |  |  | c.1490 C>T | p.Thr497Ile | Familial AF, Early PPM | Unremarkable |
| **15** | *ACTN2* |  |  | c.2161C>T | p.Arg721Cys | Unremarkable | Unremarkable |
| **16** | *CACNA1, TTN* |  |  | c.4140+5G>A, c.53969T>G | Intronic, p.Val17990Gly | SUD | Unremarkable |
| **17** | *DES* |  |  | c.727C>T | p.H243Y | SUD | LGE on CMR |
| **18** | *DES* |  |  | c.415 G>C | p.Glu139Gln | Unremarkable | Unremarkable |
| **19** | *DSP* |  |  | c.8324 C>T | p.Thr2775Ile | Familial AF | Mildly reduced LV systolic function |
| **20** | *DSP* |  |  | c.1419+5G>C | Intronic | HCM | LGE on CMR |
| **21** | *FLNC* |  |  | c.7155C>G | p.Ile2385Met | Unremarkable | Ventricular couplets on ambulatory monitor |
| **22** | *KCNH2, TTN* |  |  | c.865G>A, c.82220T>C | p.Glu289Lys, p.Ile27407Thr | SUD | LGE, NSVT on ambulatory monitor |
| **23** | *KCNJ2* |  |  | c.694 C>T | p.Leu232Phe | Familial AF | Unremarkable |
| **24** | *MYBPC3* |  |  | c.2500 C>T | p.Arg834Trp | Familial AF, Seizures, SUD | Unremarkable |
| **25** | *MYH6* |  |  | c.3808C>T | p.Arg1270Cys | Unremarkable | Ventricular couplets on ambulatory monitor |
| **26** | *MYH7* |  |  | C.3982G>A | p.Ala1328Thr | Unremarkable | Unremarkable |
| **27** | *PRKAG2* |  |  | c.1429 G>A | p.Asp477Asn | Unremarkable | Unremarkable |
| **28** | *RBM20* |  |  | c.2207A>C | p.Lys736Thr | Familial AF, SUD | LGE, NSVT on ambulatory monitor |
| **29** | *RYR2* |  |  | c.808C>T | p.His270Tyr | Unremarkable | Ventricular couplets on ambulatory monitor |
| **30** | *SCN5A* |  |  | c.1535C>T | p.Thr512Ile | Unremarkable | LGE on CMR |
| **31** | *TNNT2* |  |  | c.629A>T | p.Lys210Met | SUD | Unremarkable |
| **32** | *TPM1* |  |  | c.755G>T | p.Ser252Ile | SUD | LGE on CMR, mildly reduced LV systolic function |
| **33** | *TRPM4* |  |  | c.377G>C | p.Gly126Ala | Familial AF, ACM, SUD | Ventricular couplets on ambulatory monitor |
| **34** | *TTN* |  |  | c.60445 T>G | p.Tyr20149Asp | SUD | NSVT on ambulatory monitor |
| **35** | *TTN* |  |  | c.79547G>A | p.Gly26516Asp | Unremarkable | Unremarkable |
| **36** | *VCL* |  |  | c.622G>A | p.Ala208Thr | Familial AF | Mildly reduced LV systolic function, ventricular couplets on ambulatory monitor |

**SUPPLEMENTAL TABLE 5: Details of clinical testing**

|  | **Total**  **Cohort**  **(N=246*)** | **Genetic Evaluation Groups** | | |
| --- | --- | --- | --- | --- |
|  |  | **Positive**  **Genetic Testing**  **(N=52)** | **Gene Elusive AF Overlap**  **Syndrome**  **(N=22)** | **Negative**  **Genetic Evaluation**  **(N=172)** |
| 12-lead ECG at enrollment | 246 (100%) | 52 (100%) | 22 (100%) | 172 (100%) |
| Ambulatory monitor  None  24-48-hour Holter  7 to 30-day event monitor  Implanted device (ILR, PPM, ICD) | 37 (15.0%)  153 (62.2%)  49 (19.9%)  7 (2.8%) | 8 (15.4%)  36 (69.2%)  7 (13.5%)  1 (1.9%) | 6 (27.3%)  10 (45.5%)  5 (22.7%)  1 (4.5%) | 23 (13.4%)  107 (62.2%)  37 (21.5%)  5 (2.9%) |
| Cardiac imaging*  None  Cardiac MRI  Transthoracic echocardiogram | 16 (6.5%)  172 (69.9%)  154 (62.6%) | 2 (3.8%)  42 (80.8%)  33 (63.5%) | 1 (4.5%)  18 (81.8%)  15 (68.2%) | 13 (7.6%)  112 (65.1%)  106 (61.6%) |
| Exercise treadmill ECG | 112 (45.5%) | 34 (65.4%) | 5 (22.7%) | 73 (42.4%) |
| Sodium channel blocker challenge (procainamide) | 6 (2.4%) | 2 (3.8%) | 1 (4.5%) | 3 (1.7%) |
| *Participants may have had both a cardiac MRI and transthoracic echocardiogram | | | | |

**Supplemental Table 6: Details of phenotype penetrance and changes to management**

|  | **Overall**  **(N=246)** | **Positive Genetic Testing**  **(N=52)** | **Gene-Elusive Overlap**  **(N=22)** |
| --- | --- | --- | --- |
| **Penetrance by Gene** | | | |
| Arrhythmogenic cardiomyopathy genes |  |  |  |
| Phenotype positive for ACM/NDLVC |  | 8 (89%) |  |
| Isolated AF |  | 1 (11%) |  |
| Dilated cardiomyopathy genes |  |  |  |
| Phenotype positive for DCM |  | 13 (54%) |  |
| Isolated AF |  | 11 (46%) |  |
| Hypertrophic cardiomyopathy genes |  |  |  |
| Phenotype positive for HCM |  | 2 (40%) |  |
| Isolated AF |  | 3 (60%) |  |
| Channelopathy genes |  |  |  |
| Phenotype positive for LQTS/Brugada/PCCD/CPVT |  | 2 (50%) |  |
| Isolated AF |  | 2 (50%) |  |
| Hemochromatosis (*HFE*) |  |  |  |
| Phenotype positive for Hemochromatosis |  | 2 (50%) |  |
| Isolated AF |  | 2 (50%) |  |
| Transthyretin amyloid (*TTR*) |  |  |  |
| Phenotype positive for Amyloidosis |  | 0 (0%) |  |
| Isolated AF |  | 2 (100%) |  |
| Genes with other predominant phenotype |  |  |  |
| Other phenotype positive |  | 2 (50%) |  |
| Isolated AF |  | 2 (50%) |  |
| **Management Changes** | | | |
| Any change to clinical management following AF Precision Medicine Clinic evaluation | 37 (15.0%) | 27 (51.9%) | 7 (31.8%) |
| Anticoagulation for stroke prophylaxis started | 7 (2.8%) | 4 (7.7%) | 3 (13.6%) |
| Beta-blocker started | 21 (8.5%) | 13 (25.0%) | 6 (27.3%) |
| ACEi/ARB started | 9 (3.7%) | 6 (11.5%) | 3 (13.6%) |
| Mineralocorticoid started | 3 (1.2%) | 3 (5.8%) | 0 (0%) |
| Neprilysin inhibitor started | 4 (1.6%) | 3 (5.8%) | 1 (4.5%) |
| SGLT2 inhibitor started | 7 (2.8%) | 6 (11.5%) | 1 (4.5%) |
| Physical activity recommendations | 9 (3.7%) | 7 (13.5%) | 1 (4.5%) |
| Antiarrhythmic drug precautions | 5 (2.0%) | 5 (9.6%) | 0 (0%) |
| New permanent pacemaker | 3 (1.2%) | 3 (5.8%) | 0 (0%) |
| New implantable cardioverter defibrillator | 7 (2.8%) | 7 (13.5%) | 0 (0%) |
| New implantable loop recorder (ILR) | 3 (1.2%) | 1 (1.9%) | 1 (4.5%) |
| EP study to evaluate VT/VF risk | 4 (1.6%) | 3 (5.8%) | 1 (4.5%) |
| Therapeutic phlebotomy started | 1 (0.4%) | 1 (1.9%) | 0 (0%) |
| For TTR amyloid, Tafamadis started | 0 (0%) | 0 (0%) | 0 (0%) |
| Cascade testing performed | 56 (22.8%) | 44 (84.6%) | 1 (4.5%) |
